# Enhanced testing can substantially improve defence against several types of respiratory virus pandemic

**DOI:** 10.1101/2024.02.11.24302649

**Authors:** James Petrie, James A. Hay, Oraya Srimokla, Jasmina Panovska-Griffiths, Charles Whittaker, Joanna Masel

## Abstract

Mass testing to identify and isolate infected individuals is a promising approach for reducing harm from the next acute respiratory virus pandemic. It offers the prospect of averting hospitalizations and deaths whilst avoiding the need for indiscriminate social distancing measures. To understand scenarios where mass testing might or might not be a viable intervention, here we modelled how effectiveness depends both on characteristics of the pathogen (*R*_0_, time to peak viral load) and on the testing strategy (limit of detection, testing frequency, test turnaround time, adherence). We base time-dependent test sensitivity and time-dependent infectiousness on an underlying viral load trajectory model. We show that given moderately high public adherence, frequent testing can prevent as many transmissions as more costly interventions such as school or business closures. With very high adherence and fast, frequent, and sensitive testing, we show that most respiratory virus pandemics could be controlled with mass testing alone.

## 1 Introduction

Respiratory virus pandemics pose a major threat to human health and well-being, as evidenced by the impacts of COVID-19 and 1918 influenza pandemics. The UK government estimates between 5-25% chance of a new pandemic the magnitude of COVID-19 occurring within the next 5 years, which could potentially lead to up to 800,000 deaths as well as extensive social distancing [1]. Given the massive potential harm of future pandemics, it is important to make disease control tools more reliable. Vaccines and therapeutics currently take months or years to design, test, and manufacture, and are not guaranteed to work. Social distancing imposes a huge burden on the population and is not sustainable for a long period. With low enough disease prevalence, contact tracing can prevent transmissions more efficiently than social distancing [2]. However, contact tracing is often not reliable enough, on its own, to control airborne pathogens, especially for the short latent periods combined with pre-symptomatic and/or asymptomatic transmission seen for SARS-CoV-2 [3, 4]. Before there is another major pandemic, there is time to prepare strategies, technology, and infrastructure. It is important to understand which approaches would be most useful to invest in.

Mass testing, defined as frequent testing of most of the population and isolation of positives, was proposed as a promising approach to contain SARS-CoV-2 [5, 6, 7, 8, 9, 10, 11, 12]. On a relatively small scale (tens of thousands of people), several universities, professional sports teams and film studios succeeded in operating in-person with low disease burden during the COVID-19 pandemic by frequently testing people [13, 14, 15, 16]. Children in secondary school in the UK were recommended to test twice weekly in March 2021 [17], and Slovakia performed two rounds of mass antigen testing, which Pavelka et al [18] estimated caused a 70% reduction in prevalence. However, prior to the availability of vaccines, and even with the pooling of samples, there was not enough testing capacity outside of China [19] to consistently test more than 0.1% of the nation-wide population per day [20].

Mass testing of an entire population would be logistically complicated, and expensive, raising the question of whether scaling it up would be worth it, even in the light of the past success of smaller scale implementations. While scaling up is difficult to do in real time during a pandemic, sufficient preparation might mitigate current limitations to mass testing. Appendix A discusses how the development of technology and infrastructure might optimize for scalability, cost effectiveness, and speed.

Our primary aim is to provide a framework for understanding the potential effectiveness of different mass testing strategies (similar to Fraser et al.’s 2004 work on contact tracing [4]). If mass testing is likely to be effective, this, then this effectiveness should be weighed against expected costs to inform investments into technology and infrastructure intended to increase capacity beyond that available for SARS-CoV-2. We consider the effectiveness of mass testing given a range of likely values for viral characteristics (*R*_0_ and generation time), as well as for testing policy characteristics (limit of detection, test delay, test frequency, and population adherence). Similar to previous studies [21, 22, 23], we evaluate the potential impact of mass testing on reducing transmission, by modeling different mass testing strategies with time-dependent test sensitivity conditional on viral load trajectories. Like Middleton and Larremore [24], we treat infectiousness as a function of the same underlying viral load trajectories. In contrast to previous work, our aim is to determine which pandemic scenarios mass testing would be useful for, rather than whether it would work specifically for SARS-CoV-2. Our model is available as an interactive application (https://frequent-testing.shinyapps.io/shinyapp), to enable easy exploration of different mass testing policies and modelling assumptions.

## 2 Methods

We start by computing the expected number of transmissions from a person who tests regularly and isolates perfectly when they learn they are positive. We later adjust this value to account for imperfect adherence, and then estimate the population-level effect of mass testing under the assumption that the population is well-mixed. Default parameter values are chosen based on SARS-CoV-2, and then we explore different parameter values to represent other pathogens.

### 2.1 Viral Load Trajectory

Trajectories of log viral load for acute respiratory infections like influenza or SARS-CoV-2 are well modelled by piece-wise linear curves [15, 25, 22] (Figure 1A). For simplicity, we neglect within-population diversity in trajectories; in Appendix B we demonstrate that this assumption does not substantially change the predicted effectiveness of PCR testing against most pathogens (due to approximate linearity in relevant parts of the parameter space shown in Figure 6). This allows us to characterize pathogens according to the mean time from infection to reach peak viral load *τ*_*p*_, the mean time from peak viral load to recovery, *τ*_*r*_, and the peak viral load, *V*_*p*_. *V*_0_, the initial viral load when infected, is set to 3*·*10^−3^ based on initial infection with one viral particle and roughly 300ml of respiratory fluid [26]. Log viral load *V* is then:

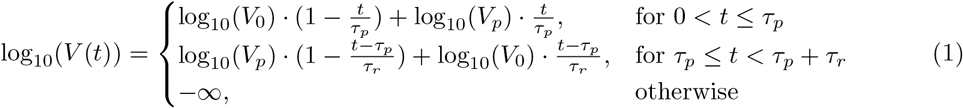

For most of this analysis, we assume viral load trajectories are symmetric, i.e. *τ*_*p*_ = *τ*_*r*_; alternative trajectory shapes can be explored further in the interactive app at https://frequent-testing.shinyapps.io/shinyapp. This assumption of symmetric trajectories slightly underestimates the effectiveness of mass testing compared to right skewed trajectories (*τ*_*r*_ *> τ*_*p*_), where transmissions occur later in infection and are therefore easier to prevent.

**Figure 1.**
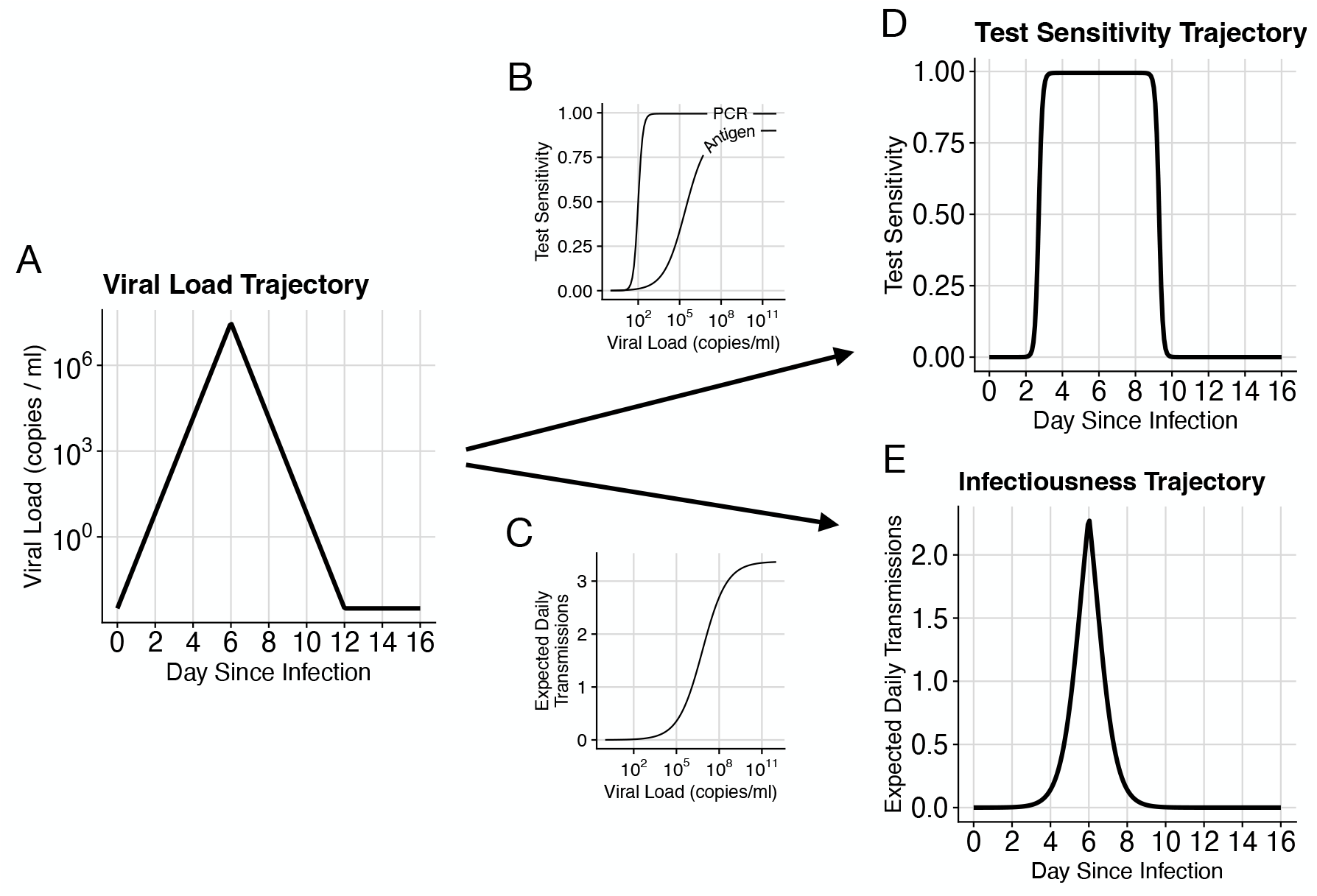
Infectiousness and test sensitivity over time are estimated based on simulated viral load trajectories. (A) An example viral load trajectory, characterized by peak viral load and times from infection to peak, and from peak to recovery. (B) Test sensitivity vs. viral load for typical PCR and antigen tests. (C) Expected transmissions per day vs. viral load. (D) Test sensitivity vs. time since infection for this pathogen, computed by passing the time series in (A) through function (B). (E) Expected daily transmissions vs. time since infection, computed by passing the time series in (A) through function (C). The area under the curve in (E) is *R*_0_ for this pathogen.

Note that inactive virus material can persist for a long time following clearance of replication competent virions, which can prolong the time from peak viral load to a negative test result. This process might therefore be better modeled as the sum of exponential declines rather than a single exponential decline. In this context, our piecewise curves can be interpreted as replication-competent viral load peaking at *V*_*P*_. Test sensitivity (probability that a sample from an infected person is classified as positive) depends strongly on the person’s viral load at the time the sample is collected [27]. We neglect the contribution to test sensitivity from non-replication competent viral material, because its only effect is to catch infected individuals too late to prevent transmission, in particular when testing is less frequent.

**Table 1:** Parameters and variables for modelling viral load, test sensitivity, and expected transmissions.

| Model Component | Description |
| --- | --- |
| $V(t; V_0, V_p, \tau_p, \tau_r)$ | Viral load at time $t$ since infection. Dependence on other parameters is described in Equation 1 and shown in Figure 1A. |
| $V_0$ | Viral load at the time of infection: $V_0 = 3 \cdot 10^{-3}$ based on one viral particle within $\approx 300$ml of respiratory fluid [26]. |
| $V_p$ | Peak viral load. Adjusted to attain a specified value of $R_0$. |
| $\tau_p$ | Time from infection to peak viral load: 1.5 to 12 days. |
| $\tau_r$ | Time from $V_p$ to $V_0$. Symmetry assumption ($\tau_r = \tau_p$) can be relaxed in the app. |
| $S(v; S_{max}, k, LOD_{50})$ | Test sensitivity (probability that a positive sample is detected as positive) as a function of viral load, $v$. Dependence on other parameters described in Equation 2 and shown in Figure 1B. |
| $S_{max}$ | Maximum test sensitivity: 99.5% for PCR, 90% for antigen tests. |
| $k$ | Controls width of intermediate test sensitivity region: $k = 6$ for PCR and $k = 1.3$ for antigen tests. |
| $LOD_{50}$ | Viral load at which test sensitivity reaches $S_{max}/2$: $10^2$ copies/ml for PCR and $10^{5.4}$ for antigen tests. |
| $T(v; N_C, h, K_m, \theta)$ | Expected transmissions per day as a function of viral load, $v$. Dependence on other parameters described by Equation 3 and shown in Figure 1C. |
| $N_C$ | Average number of close contacts per day: $N_C = 13$ based on [28]. Appendix C explores time-dependent reduction in contacts caused by symptoms. |
| $h$ | Controls width of sigmoid for transmission risk vs. viral load: $h = 0.51$ based on [29]. |
| $K_m$ | Midpoint of infectiousness: $K_m = 8.9 \cdot 10^6$ copies/ml based on [29]. |
| $\theta$ | Increased from 0.2 in Ke et al. [29] to 0.3, corresponding to a maximum per-contact transmission probability of 26%. |
| $\sigma(\delta, \rho)$ | The fraction of transmissions that would be prevented (relative to $R_0$) if the entire population tested every $\rho$ days and received test results after $\delta$ delay. |
| $\delta$ | Time delay before receiving a test result. 0 for antigen tests and 2+ hours for PCR. |
| $\rho$ | Time between tests. |
| $\gamma$ | Fraction of the population that adheres to regular testing. |
| $\beta$ | Fraction reduction in transmissions after a person who tests regularly learns they are positive. |
| $R_0$ | Expected transmissions per infected person with no population immunity or disease-control measures. |
| $R_e$ | Expected transmissions per infected person given interventions, with no population immunity. |

### 2.2 Test Sensitivity Depending on Viral Load

We model the dependence of test sensitivity on viral load, *v*, with the sigmoid function:

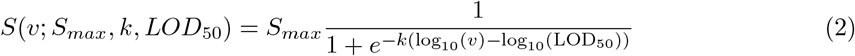

This equation is characterized by peak test sensitivity, *S*_*max*_, the viral load LOD_50_ at which *S*_*max*_*/*2 of samples from infected people are classified as positive, and a parameter *k* that specifies the width of the intermediate region. For PCR tests, we set LOD_50_ = 10^2^ copies/ml and *k* = 6 so that the distance between 5% and 95% sensitivity is about a multiple of 10 as in [27]. We set *S*_*max*_ = 99.5% to account for sample mishandling. Antigen tests are modeled as having a limit of detection of 10^5.4^ copies/ml and *k* = 1.3 based on the average sensitivity that Wagenhauser et al. measured for the ancestral SARS-CoV-2 variant [30] (although this varies considerably between manufacturers, between variants of the same pathogen, and presumably between pathogens). The maximum sensitivity, *S*_*max*_, is reduced to 90% to account for errors in self-administered antigen tests. Default PCR and antigen sensitivity curves are shown in Figure 1B.

### 2.3 Expected Transmission Rate Depending on Viral Load

While there is consensus that higher viral load increases the expected number of transmissions *T*(*v*), quantitative data on this relationship is very limited. Ke et al. [29] used the measured relationship between SARS-CoV-2 viral load (as assessed by PCR) and cell culture positivity as a proxy for the relationship between viral load and transmission. We use Ke et al.’s saturation model [29] in Equation 3, with SARS-CoV-2-inspired default values of *N*_*C*_ = 13 contacts per day [28], shape parameter *h* = 0.51, and an infectiousness midpoint of *K*_*m*_ = 8.9 *·* 10^6^copies/ml (see Appendix D for a sensitivity analysis of *K*_*m*_). Ke et al. included a parameter *θ* to reduce the maximum infectiousness below 100%; we increased this from 0.2 to 0.3, corresponding to an increase from 18% to 26% per contact.

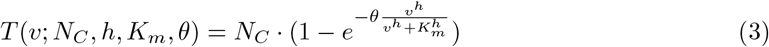

These values of *K*_*m*_ and *θ* are in broad agreement with Marc et al.’s [31] use of SARS-CoV-2 contact tracing data to connect estimated viral load at time of exposure with transmission probability. Our assessment of agreement is focused on non-household contacts, because household contacts typically are exposed over multiple days, corresponding to multiple opportunities for transmission, with a correspondingly higher overall maximum probability of infection. Equation 3 for expected daily transmissions as a function of viral load is shown in Figure 1C.

### 2.4 Test Sensitivity and Expected Transmissions over Time

Test sensitivity over time is computed as the composition of Equations 1 and 2, *S*(*V* (*t*)), shown in Figure 1D. Similarly, the expected rate of transmissions as a function of time since infection is computed as the composition of Equations 1 and 3, *T* (*V* (*t*)), shown in Figure 1E. For pathogens where noticeable symptoms cause people to reduce interactions and therefore transmit to fewer people, *N*_*C*_ can be made a function of time since infection, as explored in Appendix C (Figures 7 and 8).

### 2.5 Default Expected Transmissions per Infection (*R*_0_)

The expected number of transmissions per infected person (in an immuno-naive population with no behaviour modifications) is 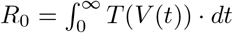 I.e., total (expected) transmission is equal to the integral of (expected) transmission over time. To achieve a given *R*_0_ value, we modify the peak viral load, *V*_*p*_, according to the relationship of Equation 1.

### 2.6 Detection Probability over Time

Consider a testing policy with a set period, *ρ*, between sequential tests. An individual whose first post-infection sample is collected *κ* days after infection (we denote this the offset) will have a negative status on day *t* following infection (i.e. will have tested negative on all samples collected on or before day *t*, if any) with probability

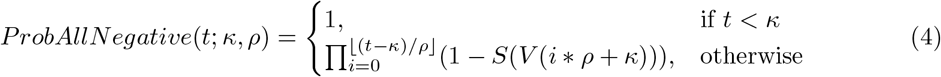

We compute the probability that a person knows they are positive at time *t* after infection by averaging across offset times, *κ ∼Uniform*(0, *ρ*) (because samples are collected every *ρ* days and infection timing is independent of test timing). Allowing for a fixed delay time, *δ*, between sample collection and receiving results, the probability that at least one sample taken before *t − δ* is detected as positive is

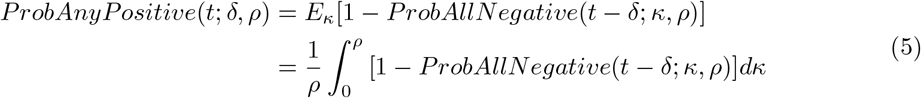

### 2.7 Transmissions Prevented by Isolation

When informing a person that they are infectious causes them to isolate effectively, the effective transmission rate at time *t* is reduced by a factor *ProbAnyPositive*(*t*). The expected number of transmissions prevented by frequent testing followed by perfect isolation after testing positive is then given by the integral of *ProbAnyPositive*(*t*) *· T* (*V* (*t*)) over time. We define *σ* as the fraction of transmissions that would be prevented by frequent testing given perfect adherence.

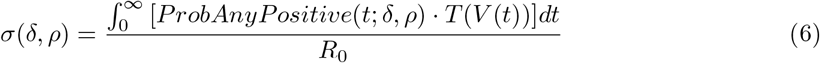

### 2.8 Transmissions with Partially Adhered to Testing and Isolation (*R*_e_)

Generalizing to imperfect adherence, let *γ* be the probability that a person adheres to regular testing, and *β* capture the degree to which a positive test results causes a person to reduce their transmission, with a fraction *σ* of their counterfactual expected transmissions having not yet occurred. Then assuming a well mixed population, mass testing reduces the effective reproduction number according to:

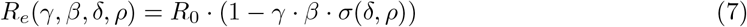

## 3 Results

Our results show that high adherence combined with frequent, rapid mass testing provide an effective strategy to prevent infections and hospitalizations. To better understand the effectiveness of mass testing, we decompose transmissions over the course of a typical infection into two broad failure modes: (1) non-adherence or (2) insufficiently sensitive, frequent, or rapid testing. Figure 2 shows expected transmissions over time when using two testing strategies against an example pathogen. Individuals that do not adhere to testing or do not effectively isolate when detected as positive, transmit on the same schedule as in the unmitigated scenario, as shown by the yellow regions in Figure 2 (the yellow regions in A and B are identical because adherence is the same for both strategies). If testing is sufficiently frequent and results are not substantially delayed, then individuals adhering to the policy transmit at a much lower rate. In this case, transmissions from adherent individuals are usually earlier in infection, as shown by the purple region in Figure 2B. When testing is more delayed (in this example 24 hours instead of 12 hours) and less frequent (here every 4 days instead of every 2 days), then adherent individuals transmit substantially more and relatively later in infection, as shown by the larger purple region in Figure 2A.

**Figure 2.**
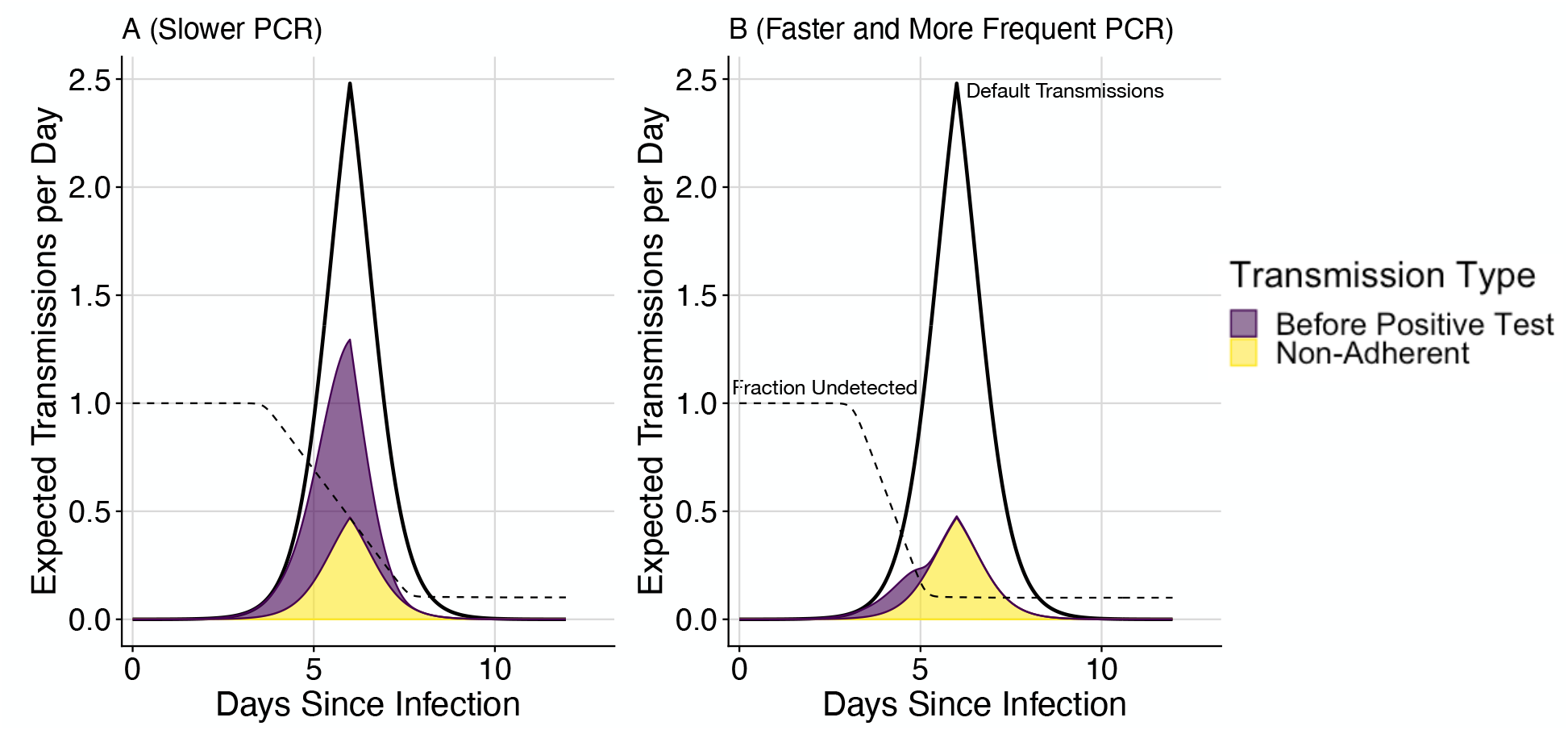
Expected transmissions under different testing strategies. The upper black lines illustrate the baseline scenario (with *R*_0_ = 4.0 expected transmissions). With mass testing, the purple areas show transmissions occurring before receiving a positive test result, while the non-overlapping yellow areas show transmissions occurring from people who don’t adhere to either testing or isolation. Dashed lines show the fraction of infected people who have not yet tested positive. A) Testing every 4 days with a 24 hour test turnaround. B) Testing every 2 days with a 12 hour test turnaround. With less frequent and more delayed tests (A), transmission reduction is modest. With more frequent and less delayed tests (B), most transmissions are prevented, and the remaining transmissions are either early in infection or due to non-adherence to the testing and isolation policy. Both scenarios use tests with a limit of detection of 10^2^ copies/ml (as described by Equation 2), 90% adherence to testing, 90% effective isolation (conditional on being a person who gets tested), 6 days from infection to peak viral load, and 6 days from peak viral load to recovery.

Figure 3 compares mass testing to other commonly used strategies against the ancestral variant of SARS-CoV-2, expressing results as transmitted averted (reduction in *R*_*e*_) as a function of adherence to testing, effectiveness of isolation, the frequency of testing, and the type of test. The proportion of transmissions prevented depends linearly on adherence to testing and isolation (Figure 3A), as described by the dependence on *γ* and *β* in Equation 7. The benefit from frequent testing saturates when testing more than every 2 days for the ancestral variant of SARS-CoV-2 (Figure 3B), with 70% more infections averted when testing every 2 days compared to every 4 days. Fast PCR testing every 3 days with 50% adherence to testing and 95% effective isolation (Figure 3B) achieves the same fraction reduction in transmissions as school and university closures (38%, as assessed by Brauner et al. [32]). Higher adherence with more frequent testing is able to further reduce transmissions (curves in figure 3B above upper dashed line).

**Figure 3.**
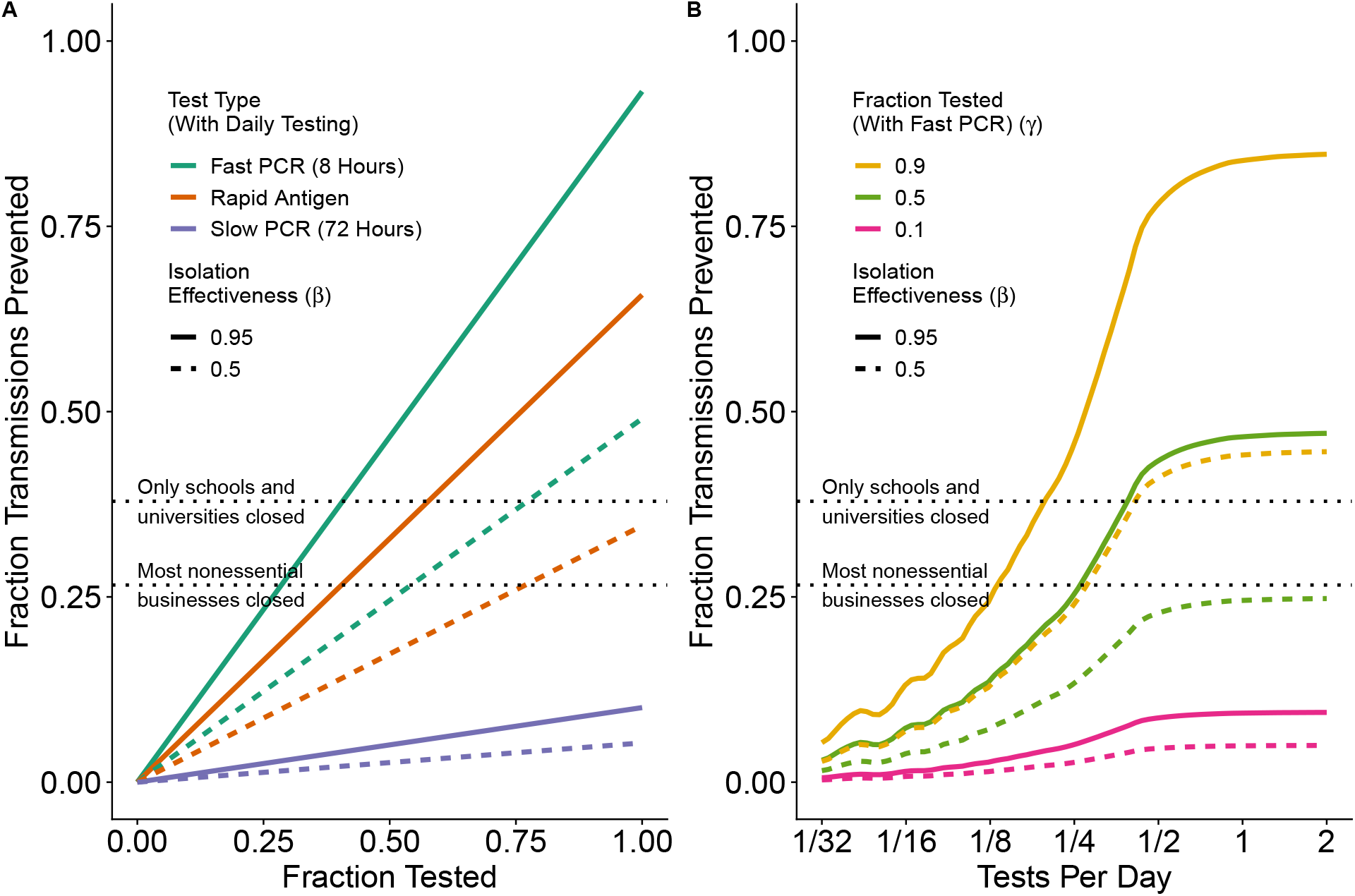
Fraction of (ancestral variant) SARS-CoV-2 transmissions prevented. depends on adherence to testing and isolation (A) and test frequency (B). SARS-CoV-2 is parameterized as taking 5 days to reach peak viral load, symptoms occurring on the same day as peak viral load, a 50% reduction in transmissions after symptoms, and *R*_0_ = 2.75 (including the behaviour modification from symptoms). In (A) we model daily testing with either a rapid antigen test (0 hour delay, *LOD*_50_ = 10^5.4^, orange), or a fast PCR test (8 hour delay, *LOD*_50_ = 10^2^, green), or a slow PCR test (72 hour delay, *LOD*_50_ = 10^2^, blue). For (B) we model only the fast PCR test. For this pathogen, the benefit of frequent testing saturates when testing more often than every 2 days (B). Estimates by Brauner et al. [32] of the fraction reduction in transmissions from school and university closures (38%) and business closures (27%) are added for comparison (dashed lines).

Fast reporting of test results is more important for diseases that reach peak viral load quickly (e.g. influenza). In Figure 4A, the fraction reduction in transmissions for a perfectly adhering individual is shown as a function of *LOD*_50_ and test delay for three example pathogens that take 3, 6, and 9 days to reach peak viral load, with peak viral load modified so that *R*_0_ = 3 for all pathogens. The contours are much steeper for the pathogen that takes 3 days to reach peak viral load than for the other pathogens, which means that the relative importance of fast reporting is greater. When using PCR tests, the number of transmissions with a 20-hour delay compared to a 10-hour delay is 3.2x larger with 3 days to peak viral load, 2.0x larger with 6 days to peak viral load, and 1.6x larger with 9 days to peak viral load. With a low limit of detection (10^2^ copies/ml) and fast reporting (10 hours), adhered-to daily testing is able to prevent more than 92% of transmissions for a pathogen with 3 days to peak viral load, and almost 99% of transmissions for a pathogen with 9 days to peak viral load.

**Figure 4.**
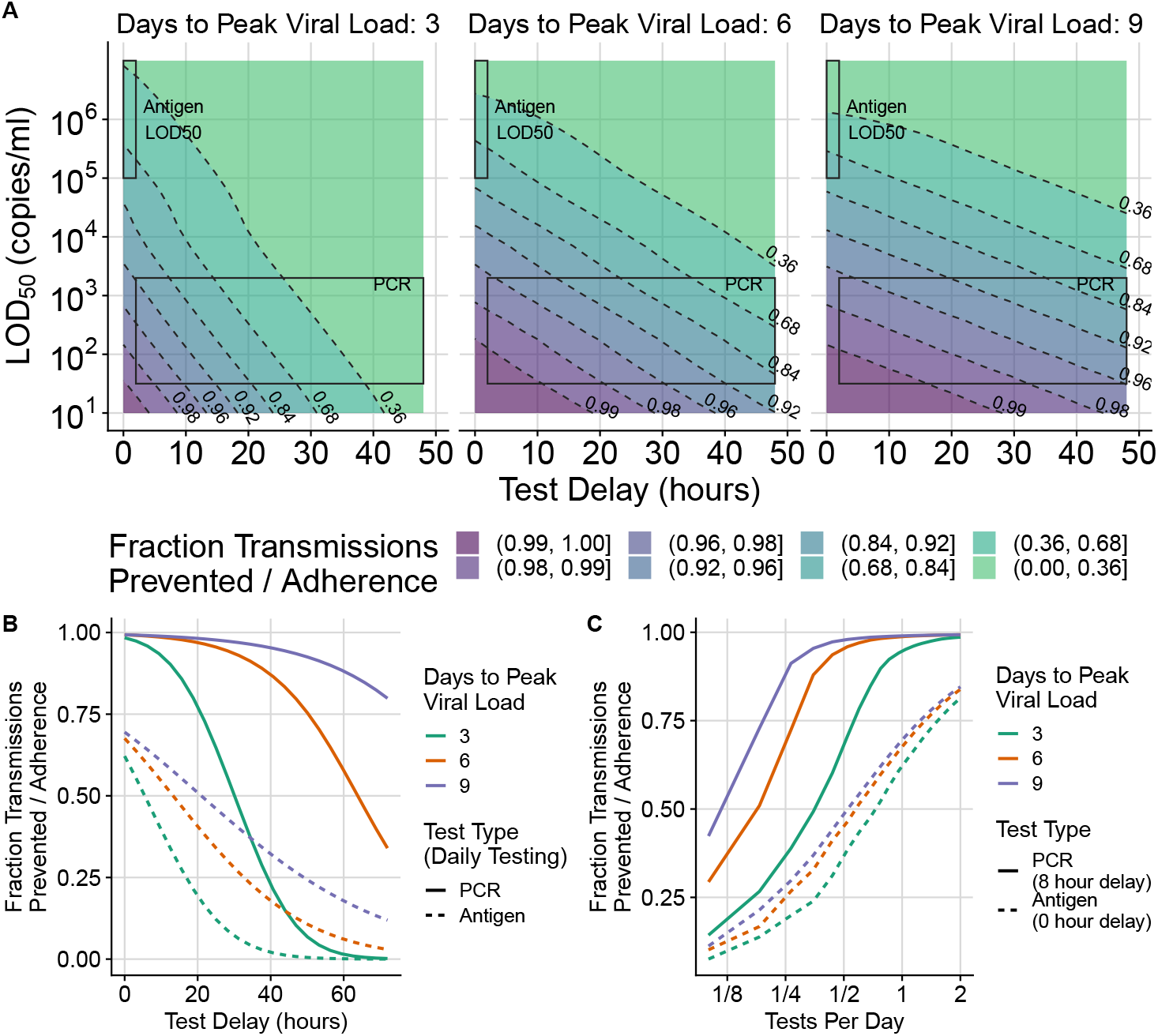
The fraction of transmissions prevented by mass testing depends on test frequency, *LOD*_50_, and test delay for three hypothetical pathogens that take 3, 6, and 9 days to reach peak viral load. The extent of transmission reduction from an infected person who tests daily and isolates effectively if positive depends on test turnaround time (x-axis of (A)), test limit of detection (y-axis of (A)), and time to peak viral load, with peak load varying accordingly to maintain *R*_0_ = 3 (3 panels in (A)). For shorter latent periods (left of (A)) delays are more important (closer contour lines along horizontal transect). Test sensitivity is similarly important for all latent periods; (similarly spaced contour lines along vertical transect). Shaded areas indicate likely *LOD*_50_ and delay time values for PCR vs. antigen tests. We use *k* = 6 and *S*_*max*_ = 99.5% from Equation 2 for all tests in (A) and for PCR tests in (B) and (C). This is overoptimistic about the maximum sensitivity (*S*_*max*_) of antigen tests, and invokes a sharper transition region than typical, so the shaded region in the upper left corner of each subplot in (A) corresponds with the *LOD*_50_ of antigen tests but not the shape of their test sensitivity curve. For (B) and (C), antigen tests are computed more accurately with *LOD*_50_ = 10^5.4^, *k* = 1.3, and *S*_*max*_ = 0.9. Rapid PCR tests (e.g. 8 hour turnaround) achieve dramatic transmission reduction under all conditions considered. Calculations assume perfect adherence to daily testing and to isolation following a positive test result. The fractional reduction in transmissions is proportional to adherence, so results can be modified for other scenarios by multiplication (e.g. if the fraction reduction is 0.8 with perfect adherence, it would be 0.4 if 50% of people tested frequently and isolated effectively if positive). Pooled PCR tests will be shifted upwards by the appropriate pooling factor, and are likely to also have increased delay. (B) shows cross-sections that hold the limit of detection constant while varying the test delay and (C) shows intervals other than daily testing.

Because of antigen tests’ higher detection limit, with *LOD*_50_ close to the viral load where someone is substantially infectious (*K*_*m*_), the predicted number of transmissions prevented is lower than for PCR. *LOD*_50_ being close to *K*_*m*_ also makes model results for antigen tests more sensitive to uncertainty in parameters, as demonstrated in Appendix D. Figure 4B shows that daily antigen testing prevents fewer transmissions than daily PCR testing unless the PCR reporting delay is more than a few days (e.g. 1 day for a pathogen with 3 days to peak viral load, and 2 days for a pathogen with 6 days to peak viral load). If antigen testing is done twice daily, Figure 4C shows that it could prevent almost 80% of transmissions; however this result should be interpreted with caution because this model assumes that subsequent test results are uncorrelated when viral load is controlled for (while in reality tests might have shared dependence on factors that might not change for a person in a day). The effectiveness of tests with different parameters can be explored using the app at https://frequent-testing.shinyapps.io/shinyapp.

To estimate the effectiveness of potential mass testing strategies against a broader range of pathogens, we compute *R*_*e*_ while varying both *R*_0_ and time to peak viral load. We find that some strategies might be able to independently control a pathogen. Using the same approach as Figure 4, we find that daily PCR testing with an 8 hour delay could avert 0.96*adherence of transmissions for a pathogen like 1918 influenza (3.5 days to peak viral load, *R*_0_ = 2.5) and 0.98*adherence for a pathogen like wild-type SARS-CoV-2 (5 days to peak viral load, *R*_0_ = 2.5). Here, adherence is defined as the product *γ · β* from Equation 7, or equivalently E[Isolation Effectiveness |Tests Regularly] ∗ P(Tests Regularly).

Mass PCR testing with overall adherence of 90% and an 8 hour reporting delay is sufficient, on its own, to control all of the example pathogens in Figure 5A except for measles (which has a very high *R*_0_). Lines in Figure 5, each representing a different testing policy, indicate the highest controllable value of *R*_0_ for each testing policy, found by increasing the peak viral load, *V*_*p*_, until *R*_*e*_ in Equation 7 is equal to 1. Figure 5A shows very high adherence (95% adherence to testing, and 95% effective isolation), while 5B shows moderately high adherence (70% adherence to testing followed by 80% isolation adherence if positive). While antigen testing in Figure 5 is insufficient on its own to control most pathogens, it can still substantially reduce the number of transmissions that are most difficult to control otherwise: transmissions from cases that have high viral load and are therefore highly contagious even during brief exposures. Using antigen tests with a lower *LOD*_50_ value can also dramatically change the results; this can be explored further at https://frequent-testing.shinyapps.io/shinyapp.

**Figure 5.**
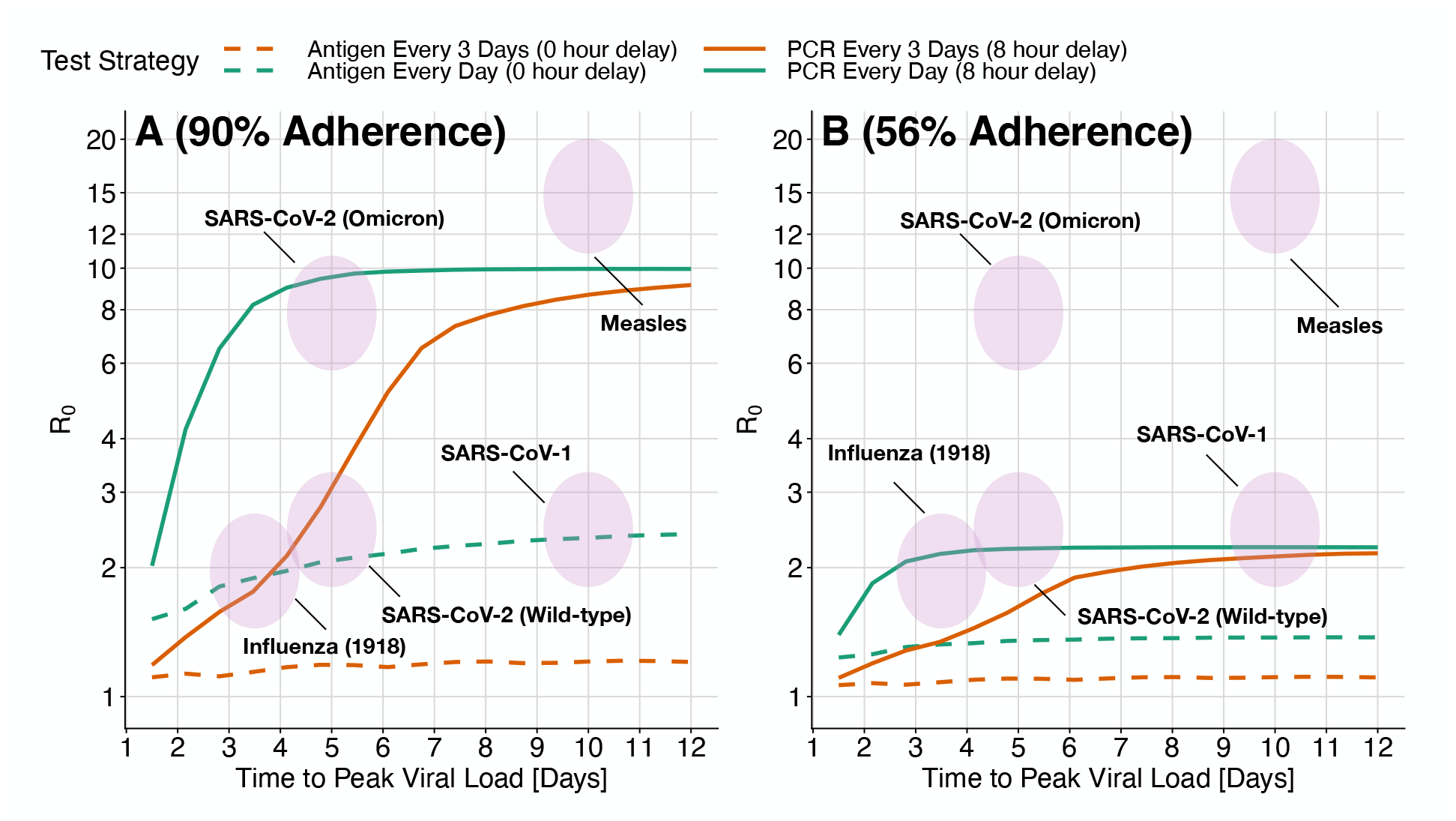
The maximum transmissibility (*R*_0_) that can be controlled by different mass testing strategies depends on the time to reach peak viral load. Viruses in the area under each line can be controlled by that testing strategy alone, in the absence of other measures. High adherence (A not B) and high test frequency (cyan) are required to control a wide variety of challenging viruses. The highest value of *R*_0_ that can be controlled with a testing strategy (computed by increasing peak viral load until *R*_*e*_ in Equation 7 is equal to 1) is shown as a function of time to peak viral load (x-axis), test type (solid vs dashed), and testing interval (orange vs cyan). Shaded ovals indicate the approximate values of *R*_0_ and time to peak viral load for a variety of viruses. Panel A) shows high adherence: 95% adherence to testing and 95% adherence to isolation if positive. The maximum *R*_0_ depends on the product of these two numbers. Panel B shows more moderate 70% adherence to testing followed by 80% isolation adherence if positive. Moderate adherence (B) might be inadequate on its own to control an outbreak of the original wild-type SARS-CoV-2 variant, but makes an important contribution when combined with other measures. As the speed of viral exponential rise and fall (x-axis) is varied, the same *R*_0_ is achieved by adjusting peak viral load. For simplicity, symptom onset does not trigger behaviour modification; in Appendix C we show similar results when symptoms are detected 24 hours before peak viral load and cause a 75% reduction in contacts (while holding *R*_0_ constant and increasing the peak viral load to compensate for fewer contacts).

While it is impossible to anticipate parameters for novel pathogens (and indeed, parameters are only partially understood for existing pathogens), the predicted effectiveness of frequent PCR testing is quite robust to changes in parameters. Different pathogens might transmit with a different proportionality factor with respect to viral load, due e.g. to how well the virus binds human cells; this will be reflected in different values of *K*_*m*_. The minimum infectious dose for SARS-CoV-2 is thought to be around 100 particles [33] and the minimum infectious dose for any pathogen has to be ≥ 1. Therefore if transmission occurs via similar mechanistic routes to SARS-CoV-2, it is unlikely that *K*_*m*_ will be more than a factor of 100 lower. In Appendix D, we explore the effect of a 100-fold lower *K*_*m*_ and show that antigen tests fail, on their own, to control any of the pathogens, while frequent PCR testing does only slightly worse (Figure 9). Similarly, the effectiveness of PCR testing is robust to behaviour modifications due to symptoms (Appendix C) and to changes in viral load trajectory shape (Viral Load tab of app: https://frequent-testing.shinyapps.io/shinyapp).

Sustained mass testing was infeasible during much of the COVID-19 pandemic, but there is now time to develop testing technology and infrastructure, so that less socially costly disease control measures are available for future pandemics. Promising newer technologies like multiplexed sequencing are becoming faster and more affordable [34]. Alternatively, more mature technology like saliva-based PCR testing can be scaled up with less technical risk, as discussed in Appendix A. In Appendix A we also discuss feasibility of potential implementations in terms of cost, public acceptability, infrastructure, and timing in a pandemic. We find that there are feasible solutions to deploy a relatively inexpensive mass testing program early in a pandemic, but that the primary logistical barrier is building and maintaining enough testing infrastructure in advance. Once built, uses will likely be found; e.g. at the time of writing, there is an acute need for more H5N1 testing of cows and farm workers in the United States[35].

## 4 Discussion

Our findings illustrate that large-scale, rapid testing can effectively control a respiratory virus pandemic, but that adherence to frequent testing and to isolation is crucial to achieve this impact. A partially adhered-to mass testing strategy could also be used to replace more restrictive interventions such as school or business closures. For the wild-type SARS-CoV-2 variant, assuming 5 days to peak viral load and *R*_0_ = 2.5, overall adherence of 61% would be needed to bring *R*_*e*_ *<* 1 in the absence of other measures. If work from home, masking, voluntary outdoor socializing etc. succeed in achieving *R*_*e*_ = 1.5, then adherence of 34% would be sufficient in a homogeneous population to further reduce *R*_*e*_ *<* 1 and achieve control. This modest level of adherence seems achievable, and could avoid the need for more draconian lockdowns. However, in scenarios where the test result delay time is long (e.g. 72 hour) compared to the time to peak viral load, mass testing becomes substantially less useful, bringing down from *R*_0_ = 2.5 to *R*_*e*_ = 2.3 and 2.4 in these two examples of 61% and 34% adherence. Contact tracing will magnify the benefits of mass testing [36], and with long test delays, the indirect benefits of testing on contact quarantine become more important than the direct effects on case isolation.

Our model assumes a well-mixed population, but we note that if there is a connected group of people that are unable or unwilling to adhere to testing or isolation, then the epidemic could escape control within that sub-network. Conversely, as seen for SARS-CoV-2 in some universities and film studios, smaller groups of people might adopt frequent testing to control transmission within their sub-network, in scenarios where there isn’t broader public support for disease elimination. The smallest, simplest example of this is testing the caregiver for someone at elevated risk of severe disease, reducing the caregiver’s transmission probability by the amount shown in Figure 4.

A scenario where massive but non-universal testing capacity could be useful is to avoid school closures by frequently testing students and staff. School closures in the US were estimated to have cost $6 trillion USD during COVID-19 pandemic, but some schools avoided closures by testing frequently. If testing were done frequently enough and a negative test were a prerequisite for attendance, then the probability of an infected contact at school transmitting the disease would be 10-20 times lower than otherwise (see Figure 4 for the 90-95% fraction reduction in transmissions, depending on the virus). Unless closing schools reduces childrens’ contacts by a factor of at least 10-20, effective testing at school could actually be safer than closing schools and not testing. This might be a good use of testing capacity that, while massive, is nevertheless insufficient on its own to control a pandemic - potentially not avoiding all social distancing measures, but avoiding school closures. While the school setting might have above the usual *R*_0_, it is also conducive to the enforcement of testing and isolation policies.

In a scenario where a pathogen causes sufficiently severe disease such that most people adopt social distancing, essential workers might choose not to work because of fear of infection. Frequent testing of essential workers and the people they interact with could dramatically improve their safety (in addition to other important safety measures like effective PPE [37]), and reduce the probability of infecting workers’ family, which could increase the number willing to continue working [38].

Frequent mass testing of asymptomatic individuals inevitably risks large numbers of false positive tests leading to isolation both of individuals who are not infected, and of individuals who were infected but are no longer infectious. However, even with wide-scale testing, the total number incorrectly isolated is manageable and fairly small, despite an expected reduction in positive predictive value as true positive individuals are removed from the population and thus prevalence in the remaining tested population goes down [39]. The duration of unnecessary isolation could be shortened by ending isolation earlier given a series of negative results, and/or by using a Ct threshold during the downward trajectory, rather than isolating until completely negative.

This work focuses on population-wide asymptomatic testing, but given limited testing capacity, it is most efficient to prioritize symptomatic testing before asymptomatic testing [40]. For simplicity, we did not model people seeking additional testing because of symptoms, although we did model behavior modification due to symptoms even in the absence of a positive test. In a related work, we use a branching model to estimate the benefit of adding symptomatic testing and contact tracing to a mass testing policy, finding that in some scenarios adding contact tracing could allow the mass testing frequency to be halved with the same reduction in transmissions [36].

We made the simplifying assumption of transmission risk being independent of location and time, but testing policies could be made more efficient by focusing on higher risk settings. For example, collecting samples from people so that their test results are returned right before attending an event would prevent even more transmissions. A location-based strategy could target more frequent testing to regions with an active outbreak, and less frequent surveillance testing for new outbreaks in previously clear regions, requiring fewer tests in total as a country approaches elimination.

The COVID-19 pandemic caused an estimated $16 trillion USD in harm in the USA [41], and efforts to contain it were also very costly (e.g. $2.5 trillion USD in lost future productivity from four months of school closure in the USA [42]). Another pandemic at least as severe as COVID-19 is likely in the next few decades, and many countries are not prepared. We show that with sufficient testing capacity and good adherence, mass testing could be effectively used to mitigate transmission and avoid the much more costly interventions and harms of future pandemics or reactions to future pandemics. This motivates substantial effort to design a testing system with high enough throughput, and to build it before the next pandemic.

## Data Availability

All code developed for this study is available online at

https://github.com/JamesPetrie/MassTestingPublic

## Acknowledgements

We would like to thank Nirav Merchant and Greg Gibson for sharing their experience with real-world testing programs, Kayla Zochowski for advice on PCR technology, Christophe Fraser for suggestions on figures, and Luca Ferretti and Chris Wymant for advice on framing. JP’s work is supported by Open Philanthropy and the Wellcome Trust (Collaboration Award 206298/Z/17/Z, ARTIC-Network). JAH is supported by a Wellcome Trust Early Career Award (grant 225001/Z/22/Z). CW is supported by Sir Henry Wellcome Postdoctoral Fellowship (reference 224190/Z/21/Z). JPG’s work is supported by the UK Health Security Agency and the UK Department of Health and Social Care. These funders had no role in the study design, data analysis, data interpretation, or writing of the report. The views expressed in this article are those of the authors and not necessarily those of the UK Health Security Agency or the UK Department of Health and Social Care.

## Appendices

### A Implementation Feasibility

For a mass testing strategy to be effective, there are several things that must go right: (1) tests must be available early in a pandemic; (2) there must be scalable logistics for fast transportation of tests/samples; (3) there must be sufficient public support for the strategy; (4) this support must be translated into sufficient adherence to testing and effective isolation; (5) there must be enough test equipment and skilled technicians; (6) the variable cost per test must be affordable. While these challenges are significant, the massive costs of either social distancing or unchecked disease, and the difficulties of effective contact tracing, motivate a thorough investigation of mass testing as a potential solution, either as an alternative to distancing and contact tracing, or as an adjunct.

In this Appendix we focus on PCR testing of saliva samples because it is a mature technology that has been proven at scale and can be functional within weeks of detecting a pandemic. Rapid antigen testing is another option that has been proven at scale, but for it to be useful in future pandemics, development, manufacturing, and deployment would have to be much faster than they were during the COVID-19 pandemic. Specifically, rapid antigen tests became available almost a year after detecting the COVID-19 pandemic [43]. There are also several other promising technologies, e.g. based on multiplexed next-generation sequencing [34, 44], LAMP [45], CRISPR [46], or even particle imaging [47]. The aim of this Appendix is to demonstrate one relatively low-risk strategy for scaling up, but not to argue that it is the best or only possible solution for mass testing. In the ideal case, redundant technological approaches and infrastructure could improve robustness to unanticipated challenges.

#### A.1 Time to Deploy

PCR can be ready within weeks of detecting a pathogen; e.g., Corman et al. published a protocol for SARS-CoV-2 on January 23, 2020, 13 days after the pathogen sequence was first published [48]. The main decision that needs to be made is which amplicon targets to use to selectively amplify RNA or DNA from the pathogen of interest (e.g. distinguishing SARS-CoV-2 from other coronaviruses) [49]. After a protocol is chosen, oligonucleotides for the target amplicons can be synthesized with a high-throughput solid phase process [50] and distributed to testing labs. Redundancy in protocol design and test kit manufacturing would be prudent to avoid delays like the ones experienced by the USA in early 2020 [51].

#### A.2 Logistics: Sample Collection and Transportation

Cost-effective deployment at scale could be achieved by borrowing approaches from home delivery services, a market that has recently expanded and successfully overcome many logistical challenges. As an example, both unused sample collection kits and self-collected saliva samples could be left at unstaffed booths (potentially with security cameras to deter misuse). This could be done cheaply by providing a tray of empty tubes from which users detach a QR code, collect and seal a saliva sample, and place the tube in a box. Rideshare or delivery companies could be hired on demand to frequently (i.e. ideally with less than 4 hours wait time to achieve a total turnaround less than 8 hours) pick up the boxes of samples at a low average cost per sample (e.g. if 50 samples were transported by a $30 USD Uber ride). In low or middle income countries (LMICs) the available budget would likely be lower, but the cost of labour would also be lower. The right choices for logistics depend on the setting, but using self-collected samples with batched delivery through existing consumer delivery services or similar could likely keep the cost of logistics below $1 per sample in high income countries and considerably lower in LMICs.

#### A.3 Public Support

Mass testing depends on public support, which could potentially be improved by making the overall approach simple and non-invasive. For example, several schools used saliva or gargle based testing because it avoids the discomfort of nasopharyngeal swabs [52] and samples are easily self-collected without specialized supplies. Saliva tests sometimes had higher sensitivity early in infection [53], although the cheap, low-volume SalivaDirect test had slightly worse sensitivity [54]. Using easily accessible locations where tests can be completed with a few minutes could also improve the overall experience. Because public support is so important for mass testing to be successful, it is likely worthwhile to make significant trade-offs in test sensitivity, delay time, or cost if necessary. However, if there remains insufficient public support for mass testing of the entire population, testing could be focused on schools and senior living facilities.

#### A.4 Testing and Isolation Adherence

The achievable level of testing and isolation adherence depends on public sentiment, which likely depends strongly on the severity of the pandemic. In milder pandemics, adherence might depend on voluntary participation, as enforcement might be too extreme an infringement on civil liberties. However, in a very deadly pandemic without viable alternative control strategies, there might be support for more strictly enforcing adherence. This might be easier if the verification process is smooth, and if those who do not wish to adhere have some viable (albeit restrictive) alternative option to partially quarantine rather than test. E.g., proof of a recent negative test could be required for entry into public spaces where there is substantial risk of transmission (similar to testing requirements for flights or some schools or workplaces for SARS-CoV-2).

Data on the effectiveness of home isolation is limited [55], but isolation in dedicated facilities is likely to be highly effective. Even quarantine in poorly chosen facilities that mixed air between the infected and uninfected was shown to be at least 99.4% effective at preventing transmissions of SARS-CoV-2 from infected cases in Australia [56]. Isolation could be incentivized by fully replacing lost wages (the COVID-19 pandemic typically saw partial wage replacement at best [57]), and providing generous support (e.g. accommodation, food, childcare, medical care) [55]. Successful local control of a pandemic corresponds to fewer than 1/1000 people infected at any time (and hopefully far fewer), meaning that spending double each person’s salary to support their isolation would cost less than 2/1000 of GDP.

#### A.5 Testing Capacity: Equipment and Skilled Labour

Test capacity depends on equipment and skilled labour. Capacity was a significant barrier to performing mass testing during the COVID-19 response when there was little time to manufacture, let alone design, new equipment. Sample pooling was sometimes used to test more people using a fixed amount of available equipment, at the expense of decreased sensitivity, increased complexity, and many re-tests when prevalence is high [58].

A simple way to build PCR capacity in preparation for future pandemics is to manufacture more conventional PCR equipment, to be combined with sample pooling. Minhas et al’s description of a national lab in Pune, India [59] can be used as a basis to estimate the upfront investment and number of trained staff needed for this strategy, although we note that locations with high labour costs would probably employ fewer people while using more expensive equipment. Their peak capacity was 1800 samples per day, they employed around 100 people, and the cost of their diagnostic equipment was 15,592,420 INR, which is roughly equivalent to $188,000 USD. The same lab could process samples that were pooled 10x at time of collection (having all 10 individuals home isolate until individually retested), thus processing roughly 18,000 samples per day. 100 people employed in a lab per 20,000 sampled is 1/200 of the total population; this would be highly costly to staff in preparation. As a comparison, 1/2000 people are on ‘standby’ as firefighters in the UK [60]. Rapidly recruiting skilled technicians when a pandemic is detected is another option, but this would be difficult to do in 1-2 months and only about 1/1000 people are currently employed in the medical diagnostics industry in the US [61].

Alternatively, there has been recent progress on highly automated workflows for PCR diagnostics [62, 63, 64] that require fewer lab technicians. One example is endpoint PCR [65], which uses a waterbath instead of the more common piezoelectric system, for higher-throughput thermal cycling at lower cost. A large British diagnostics company, LGC Group, claims to be able to test 150,000 samples per day [66] with two technicians per shift and an upfront cost of $902,000 USD for the equipment [personal communication]. We note however that a real-world deployment of this technology only reached 65,000 samples/day with an infrastructure cost of $186M USD [67]. The reasons for such a high cost include construction of a 220,000 square foot facility, while the low output likely reflects reliability issues with early-stage technology [68].

One strategy for building and maintaining high-throughput PCR for the purpose of pandemic readiness, within a developed country, is to make research institutions eligible for targeted grants. These grants would provide standardized ultra-high-throughput PCR equipment, which the grantee would be free to put to innovative research use between emergencies. In exchange, grantees would guarantee a mobilizable workforce able to use the equipment for mass testing, as assessed through a system of occasional drills. Once established, it might be found that mass testing is useful more often than anticipated, e.g. at the time of writing, there is an acute need for more H5N1 testing of cows and farm workers in the United States [35]. Such a scheme would have the benefit of standardizing the equipment, once an appropriate prototype is available. The current LGC technology would first need clearer proof of concept before being chosen, and it is possible that more R&D is first required to build an adequate system.

#### A.6 Variable Cost per Test

Even when there is sufficient equipment and trained personnel, there is a per-test cost that determines how much countries can afford to test in a pandemic. This cost depends on the price of consumables and the price of logistics (discussed in Section A.2), and was fairly high in most countries during the COVID-19 pandemic (e.g. $24 - $55 USD per test in the UK [69]). Pooled testing substantially reduced the cost of PCR testing during the COVID-19 pandemic by using less consumables per sample (e.g. Mirimus Inc. tested students in New York for $10/sample [70], and China tested millions of people for $1.50/sample [58]). SalivaDirect demonstrated that low prices could be achieved by using a small volume of the cheapest available reagents, reducing the cost of consumables to $1.21/sample with an extraction free protocol [54]. In Section A.5 we argued that the cost of logistics in high income countries could be less than $1 per sample, so with consumables included the cost per test could be around $2. For wealthier countries, a cost of $2/person/day is very affordable if it averts most of the harm of the pandemic, and for LMICs, the cost could potentially be further reduced with sample pooling and when considering the lower cost of delivery. For the USA, the cost of testing every person daily for a year would be about $240 billion, which is almost 10 times smaller than the $2.2 trillion spent on the CARES Act [71] in response to the COVID-19 pandemic.

### B Impact of Heterogeneity in Transmissibility and Time to Peak Viral Load

We modeled each pathogen as having a fixed expected number of transmissions (*R*_0_) and time to reach peak viral load (*τ*_*p*_) for every infected person. In reality, viral load trajectories will be different for every person who is infected, which could be more accurately modelled as drawing *R*_0_ and *τ*_*p*_ from a distribution. By only computing *R*_*e*_ at the average parameter values instead of integrating over the entire parameter distribution we might introduce some error. I.e., we made the approximation that *E*[*R*_*e*_(*τ*_*p*_, *R*_0_)] ≈ *R*_*e*_(*E*[*τ*_*p*_], *E*[*R*_0_]), where [*τ*_*p*_, *R*_0_] is a random vector.

If *R*_*e*_(*τ*_*p*_, *R*_0_) is a linear function, then this approximation is exact. If *R*_*e*_(*τ*_*p*_, *R*_0_) is well approximated by a linear function in the neighborhood occupied by the parameter distribution for a pathogen, then the error introduced by this approximation is therefore small. Figure 6 shows how *R*_*e*_ depends on *R*_0_ and *τ*_*p*_ for PCR tests and antigen tests (assuming perfect adherence - lower adherence can be computed with a linear transformation of the displayed function). Approximately linear regions in the function are shown as either not having any contour lines (flat) or having consistent spacing between straight contour lines (increasing linearly in the direction of a constant vector). For both types of tests the function is nonlinear when *τ*_*p*_ *<* 3 days, and for antigen tests the function becomes nonlinear with lower values of *R*_0_. If the [*τ*_*p*_, *R*_0_] distribution for a pathogen does not have much mass in these non-linear regions, then the point approximation does not cause significant error.

**Figure 6.**
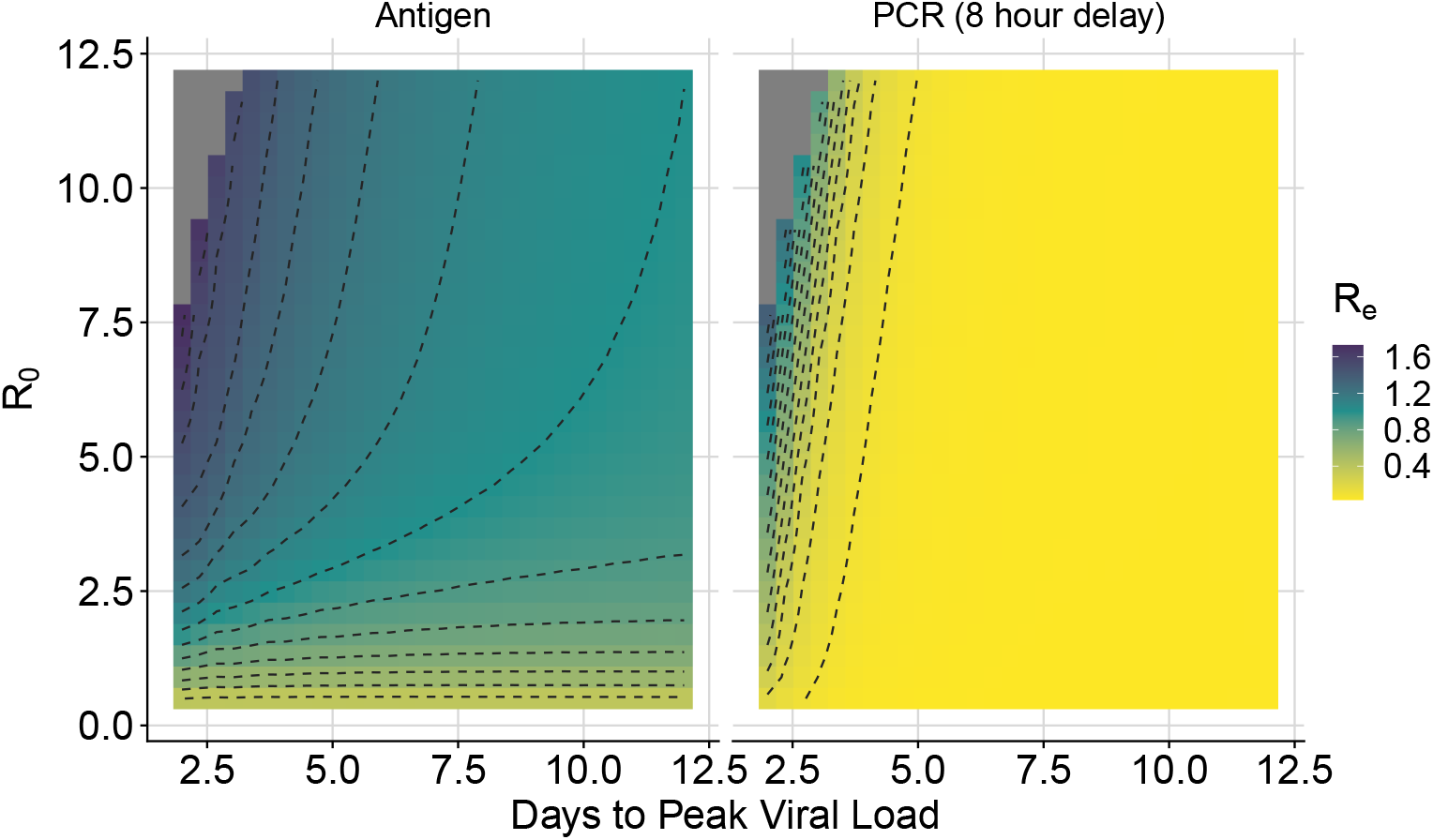
*R*_*e*_ vs. *R*_0_ and *τ*_*p*_ for daily antigen testing (with immediate results) and PCR testing (with an 8 hour result delay).

### C Sensitivity to Symptom-Based Behaviour Modification

Modified assumptions regarding timing of symptoms and behaviour change due to symptoms do not substantially change the estimated effectiveness of mass testing. Figure 7 is generated using the same parameters as Figure 3, except symptoms occur 24 hours before peak viral load instead of at the time of peak viral load, and contacts are reduced 75% instead of 50% after symptoms. Because *R*_0_ is held at the same value of 2.75, the peak viral load for the scenario with earlier symptoms is higher. The predicted impact of testing in these two scenarios is almost identical, except with earlier symptoms, slightly more frequent testing is needed. This is mainly because the increase in peak viral load shifts more transmissions earlier in the infection, which requires more frequent testing to detect in time. Similarly, Figure 8 shows a modified version of Figure 5 in the scenario where symptoms are detected 24 hours before peak viral load, and cause a 75% reduction in transmissions. For a fixed *R*_0_, the addition of symptoms changes the shape of the infectiousness profile. Daily testing performs slightly worse in this scenario because more transmissions occur earlier in infection. With testing every 3 days for fast viruses, the increase in test sensitivity from a greater viral load is a more important effect, causing the testing effectiveness to increase slightly.

**Figure 7.**
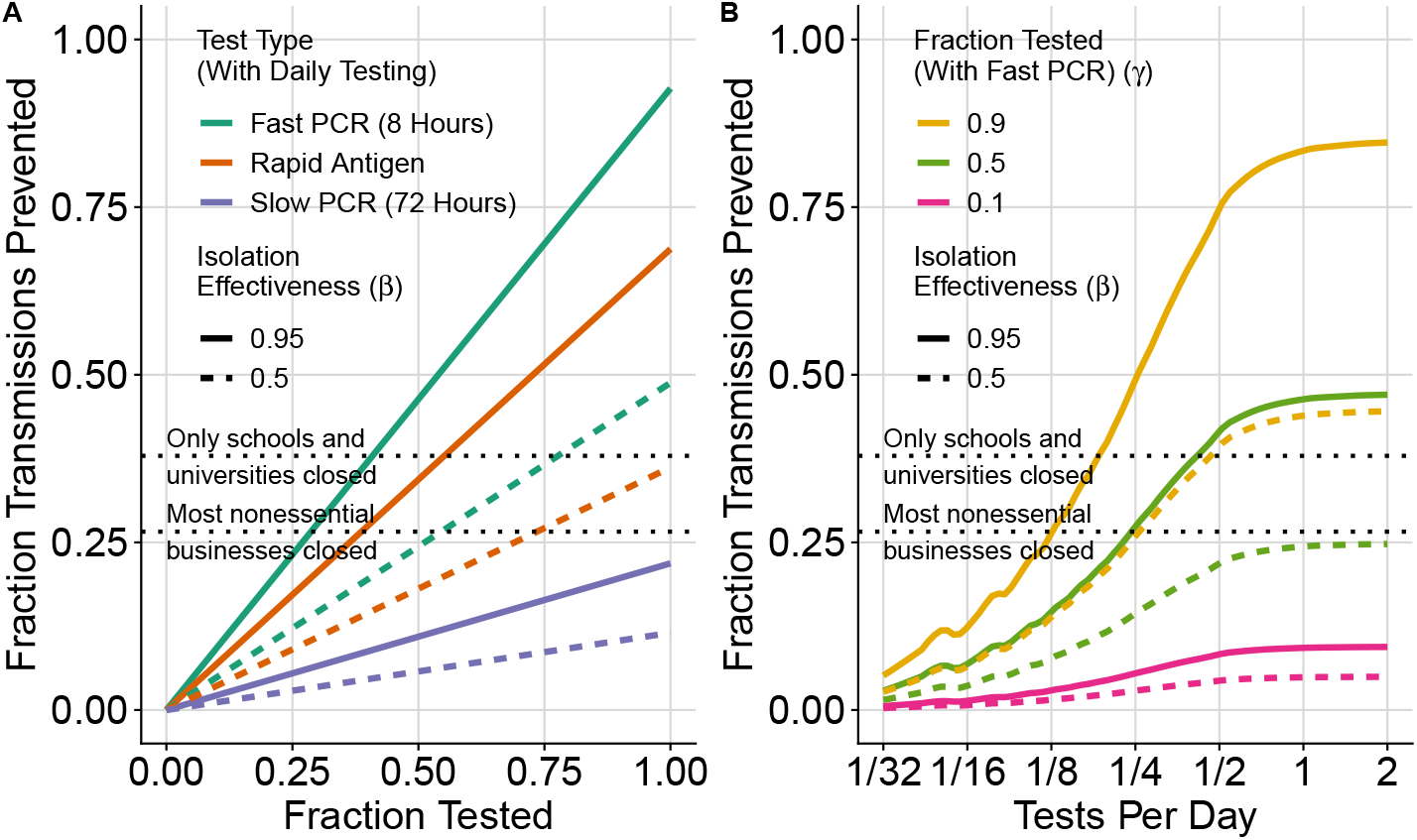
Fraction of (ancestral variant) SARS-CoV-2 transmissions prevented when symptoms occur 1 day before peak viral load, and contacts are reduced by 75% after symptoms. Compare to symptoms 0 days before peak viral load and a 50% reduction in contacts in Figure 3). Because *R*_0_ is kept at 2.75, the additional behaviour modification from symptoms causes the computed peak viral load to increase slightly, causing transmissions to shift earlier in infection. Except for a slight reduction in the effectiveness of slow PCR tests, the modified symptom parameters do not substantially change Figure 3.

**Figure 8.**
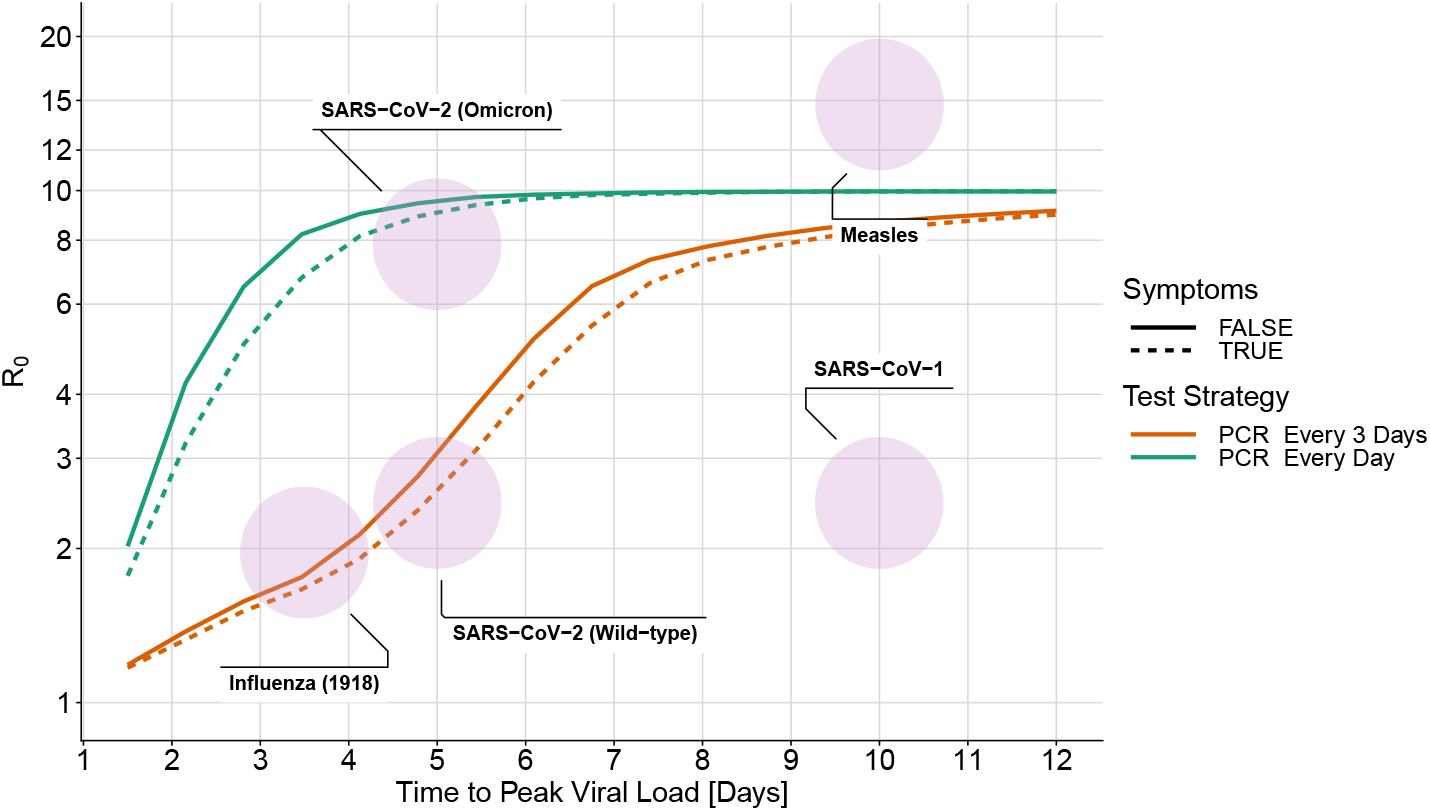
Effectiveness of mass testing with and without behaviour modification due to symptoms. As in Figure 5, the solid lines do not include symptoms. The dashed lines are computed with symptoms 24 hours before peak viral load and a 75% reduction in contacts after symptoms. For a fixed value of *R*_0_, the addition of symptoms causes the computed viral load to increase and transmissions to shift earlier in infection. Earlier transmissions are generally more difficult to control, so the effectiveness of mass testing decreases slightly because of this.

### D Sensitivity to Infectiousness Midpoint

To test the sensitivity of our results to the infectiousness function in Equation 3, we reduce the viral load at which half of peak infectiousness has been reached from *K*_*m*_ = 8.9 *·* 10^6^ to *K*_*m*_ = 8.9 *·* 10^4^. With a 100 times smaller value of *K*_*m*_, the viral load needed to transmit is substantially lower, while the viral load needed to detect infection stays the same. In Figure 9 we see that with the reduced value of *K*_*m*_, frequent PCR testing does slightly worse than in Figure 5, while antigen testing fails to control any of the pathogens. This is because in this extreme scenario, PCR tests are still able to detect infections before people become substantially infectious, while the antigen tests fail to detect infected people even at their highest viral load.

**Figure 9.**
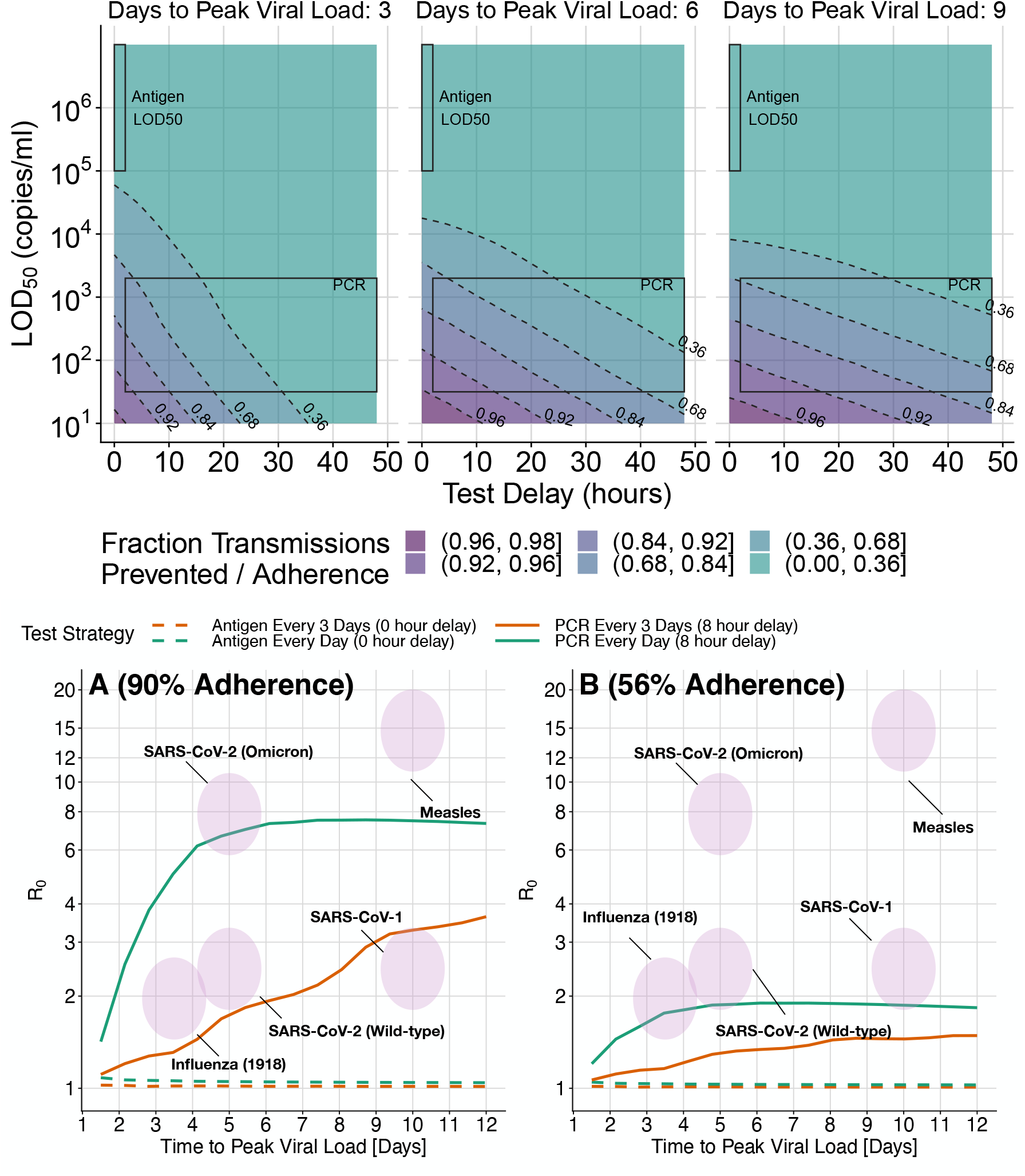
Figures 4 and 5, are recomputed with the viral load midpoint for infectiousness reduced from *K*_*m*_ = 8.9 · 10^6^ to *K*_*m*_ = 8.9 · 10^4^. In the upper 3 panels we see that for PCR tests, the number of transmissions prevented is slightly lower than in Figure 4, and for antigen tests less than 30% of the transmissions are prevented. In the bottom two panels, PCR tests are able to control a similar range of epidemics as in Figure 5 (with slightly reduced effectiveness) and antigen tests are unable to substantially control any outbreaks.

## References

1. Cabinet Office. National Risk Register 2023. Available from: https://www.gov.uk/government/publications/national-risk-register-2023 [Accessed on: 2023 Aug 10]

2. Petrie J and Masel J. Optimal targeting of interventions uses estimated risk of infectiousness to control a pandemic with minimal collateral damage. 2023 Oct. doi: 10.1101/2023.10.06.23296661. Available from: https://www.medrxiv.org/content/10.1101/2023.10.06.23296661v1 [Accessed on: 2023 Nov 20]

3. Davis EL, Lucas TCD, Borlase A, Pollington TM, Abbott S, Ayabina D, Crellen T, Hellewell J, Pi L, Medley GF, Hollingsworth TD, and Klepac P. Contact tracing is an imperfect tool for controlling COVID-19 transmission and relies on population adherence. Nature Communications 2021 Sep; 12. Number: 1 Publisher: Nature Publishing Group:5412. doi: 10.1038/s41467-021-25531-5. Available from: https://www.nature.com/articles/s41467-021-25531-5

4. Fraser C, Riley S, Anderson RM, and Ferguson NM. Factors that make an infectious disease outbreak controllable. Proceedings of the National Academy of Sciences 2004 Apr; 101. Publisher: Proceedings of the National Academy of Sciences:6146–51. doi: 10.1073/pnas.0307506101. Available from: https://www.pnas.org/doi/full/10.1073/pnas.0307506101

5. Colbourn T, Waites W, Panovska-Griffiths J, Manheim D, Sturniolo S, Colbourn G, Bowie C, Godfrey KM, Peto J, and Burgess RA. Modelling the health and economic impacts of population-wide testing, contact tracing and isolation (PTTI) strategies for COVID-19 in the UK. 2020. Available from: https://papers.ssrn.com/sol3/papers.cfm?abstract_id=3627273

6. Taipale J, Romer P, and Linnarsson S. Population-scale testing can suppress the spread of COVID-19. 2020 May. doi: 10.1101/2020.04.27.20078329. Available from: https://www.medrxiv.org/content/10.1101/2020.04.27.20078329v2 [Accessed on: 2023 Nov 2]

7. Libin PJK, Willem L, Verstraeten T, Torneri A, Vanderlocht J, and Hens N. Assessing the feasibility and effectiveness of household-pooled universal testing to control COVID-19 epidemics. PLOS Computational Biology 2021 Mar; 17. Publisher: Public Library of Science:e1008688. doi: 10.1371/journal.pcbi.1008688. Available from: https://journals.plos.org/ploscompbiol/article?id=10.1371/journal.pcbi.1008688

8. Larremore DB, Wilder B, Lester E, Shehata S, Burke JM, Hay JA, Tambe M, Mina MJ, and Parker R. Test sensitivity is secondary to frequency and turnaround time for COVID-19 screening. Science Advances 2021 Jan; 7. Publisher: American Association for the Advancement of Science:eabd5393. doi: 10.1126/sciadv.abd5393. Available from: https://www.science.org/doi/10.1126/sciadv.abd5393

9. Neilan AM, Losina E, Bangs AC, Flanagan C, Panella C, Eskibozkurt GE, Mohareb A, Hyle EP, Scott JA, Weinstein MC, Siedner MJ, Reddy KP, Harling G, Freedberg KA, Shebl FM, Kazemian P, and Ciaranello AL. Clinical Impact, Costs, and Cost-effectiveness of Expanded Severe Acute Respiratory Syndrome Coronavirus 2 Testing in Massachusetts. Clinical Infectious Diseases: An Official Publication of the Infectious Diseases Society of America 2020 Sep; 73:e2908–e2917. doi: 10.1093/cid/ciaa1418. Available from: https://www.ncbi.nlm.nih.gov/pmc/articles/PMC7543346/

10. Poole SF, Gronsbell J, Winter D, Nickels S, Levy R, Fu B, Burq M, Saeb S, Edwards MD, Behr MK, Kumaresan V, Macalalad AR, Shah S, Prevost M, Snoad N, Brenner MP, Myers LJ, Varghese P, Califf RM, Washington V, Lee VS, and Fromer M. A holistic approach for suppression of COVID-19 spread in workplaces and universities. PLOS ONE 2021 Aug; 16. Publisher: Public Library of Science:e0254798. doi: 10.1371/journal.pone.0254798. Available from: https://journals.plos.org/plosone/article?id=10.1371/journal.pone.0254798

11. Lehrach H, Curtis J, Lange B, Ogilvie LA, Gauss R, Steininger C, Scholz E, and Kreck M. Proposal of a population wide genome-based testing for Covid-19. Scientific Reports 2022 Apr; 12. Number: 1 Publisher: Nature Publishing Group:5618. doi: 10.1038/s41598-022-08934-2. Available from: https://www.nature.com/articles/s41598-022-08934-2

12. Cherif R and Hasanov F. Testing our way out of pandemics. Health Policy and Technology 2023 Mar; 12:100714. doi: 10.1016/j.hlpt.2022.100714. Available from: https://www.sciencedirect.com/science/article/pii/S2211883722001216

13. Ranoa DRE et al. Mitigation of SARS-CoV-2 transmission at a large public university. Nature Communications 2022 Jun; 13. Number: 1 Publisher: Nature Publishing Group:3207. doi: 10.1038/s41467-022-30833-3. Available from: https://www.nature.com/articles/s41467-022-30833-3

14. Gibson G, Weitz JS, Shannon MP, Holton B, Bryksin A, Liu B, Sieglinger M, Coenen AR, Zhao C, Beckett SJ, Bramblett S, Williamson J, Farrell M, Ortiz A, Abdallah CT, and García AJ. Surveillance-to-Diagnostic Testing Program for Asymptomatic SARS-CoV-2 Infections on a Large, Urban Campus in Fall 2020. Epidemiology 2021 Nov; 33:209–16. doi: 10.1097/EDE.0000000000001448

15. Kissler SM, Fauver JR, Mack C, Olesen SW, Tai C, Shiue KY, Kalinich CC, Jednak S, Ott IM, Vogels CBF, Wohlgemuth J, Weisberger J, DiFiori J, Anderson DJ, Mancell J, Ho DD, Grubaugh ND, and Grad YH. Viral dynamics of acute SARS-CoV-2 infection and applications to diagnostic and public health strategies. PLOS Biology 2021 Jul; 19. Publisher: Public Library of Science:e3001333. doi: 10.1371/journal.pbio.3001333. Available from: https://journals.plos.org/plosbiology/article?id=10.1371/journal.pbio.3001333

16. Rossol M. Comparison of Two Entertainment Industry COVID-19 Programs. NEW SOLUTIONS: A Journal of Environmental and Occupational Health Policy 2022 Nov; 32. Publisher: SAGE Publications Inc:171–81. doi: 10.1177/10482911221134515. Available from: 10.1177/10482911221134515

17. Hughes DM, Bird SM, Cheyne CP, Ashton M, Campbell MC, García-Fiñana M, and Buchan Rapid antigen testing in COVID-19 management for school-aged children: an observational study in Cheshire and Merseyside, UK. Journal of Public Health 2023 Mar; 45:e38–e47. doi: 10.1093/pubmed/fdac003. Available from: 10.1093/pubmed/fdac003

18. Pavelka M, Van-Zandvoort K, Abbott S, Sherratt K, Majdan M, CMMID COVID-19 WORKING GROUP, InšTitúT ZdravotnýCh AnalýZ, Jarčuška P, Krajčí M, Flasche S, and Funk S. The impact of population-wide rapid antigen testing on SARS-CoV-2 prevalence in Slovakia. Science 2021 May; 372. Publisher: American Association for the Advancement of Science:635–41: 10.1126/science.abf9648. Available from: https://www.science.org/doi/10.1126/science.abf9648

19. Zhou Y, Jiang H, Wang Q, Yang M, Chen Y, and Jiang Q. Use of contact tracing, isolation, and mass testing to control transmission of covid-19 in China. BMJ 2021 Dec; 375. Publisher: British Medical Journal Publishing Group Section: Analysis:n2330. doi: 10.1136/bmj.n2330. Available from: https://www.bmj.com/content/375/bmj.n2330

20. Daily COVID-19 tests per 1,000 people. Available from: https://ourworldindata.org/grapher/daily-tests-per-thousand-people-smoothed-7-day [Accessed on: 2023 Aug 12]

21. Brault V, Mallein B, and Rupprecht JF. Group testing as a strategy for COVID-19 epidemiological monitoring and community surveillance. PLOS Computational Biology 2021 Mar; 17. Publisher: Public Library of Science:e1008726. doi: 10.1371/journal.pcbi.1008726. Available from: https://journals.plos.org/ploscompbiol/article?id=10.1371/journal.pcbi.1008726

22. Cleary B, Hay JA, Blumenstiel B, Harden M, Cipicchio M, Bezney J, Simonton B, Hong D, Senghore M, Sesay AK, Gabriel S, Regev A, and Mina MJ. Using viral load and epidemic dynamics to optimize pooled testing in resource-constrained settings. Science Translational Medicine 2021 Apr; 13. Publisher: American Association for the Advancement of Science:eabf1568. doi: 10.1126/scitranslmed.abf1568. Available from: https://www.science.org/doi/10.1126/scitranslmed.abf1568

23. Yu J, Huang Y, and Shen ZJ. Optimizing and evaluating PCR-based pooled screening during COVID-19 pandemics. Scientific Reports 2021 Nov; 11. Number: 1 Publisher: Nature Publishing Group:21460. doi: 10.1038/s41598-021-01065-0. Available from: https://www.nature.com/articles/s41598-021-01065-0

24. Middleton C and Larremore DB. Modeling the Transmission Mitigation Impact of Testing for Infectious Diseases. 2023 Sep. doi: 10.1101/2023.09.22.23295983. Available from: https://www.medrxiv.org/content/10.1101/2023.09.22.23295983v1 [Accessed on: 2023 Oct 24]

25. Carrat F, Vergu E, Ferguson NM, Lemaitre M, Cauchemez S, Leach S, and Valleron AJ. Time Lines of Infection and Disease in Human Influenza: A Review of Volunteer Challenge Studies. American Journal of Epidemiology 2008 Apr; 167:775–85. doi: 10.1093/aje/kwm375. Available from: 10.1093/aje/kwm375

26. Lewis FR, Elings VB, and Sturm JA. Bedside measurement of lung water. Journal of Surgical Research 1979 Oct; 27:250–61. doi: 10.1016/0022-4804(79)90138-0. Available from: https://www.sciencedirect.com/science/article/pii/0022480479901380

27. Yang J, Han Y, Zhang R, Zhang R, and Li J. Comparison of analytical sensitivity of SARS-CoV-2 molecular detection kits. International Journal of Infectious Diseases 2021 Oct; 111:233–41. doi: 10.1016/j.ijid.2021.08.043. Available from: 10.1016/j.ijid.2021.08.043

28. Mossong J, Hens N, Jit M, Beutels P, Auranen K, Mikolajczyk R, Massari M, Salmaso S, Tomba GS, Wallinga J, Heijne J, Sadkowska-Todys M, Rosinska M, and Edmunds WJ. Social Contacts and Mixing Patterns Relevant to the Spread of Infectious Diseases. PLOS Medicine 2008 Mar; 5. Publisher: Public Library of Science:e74. doi: 10.1371/journal.pmed.0050074. Available from: https://journals.plos.org/plosmedicine/article?id=10.1371/journal.pmed.0050074

29. Ke R, Zitzmann C, Ho DD, Ribeiro RM, and Perelson AS. In vivo kinetics of SARS-CoV-2 infection and its relationship with a person’s infectiousness. Proceedings of the National Academy of Sciences 2021 Dec; 118. Publisher: Proceedings of the National Academy of Sciences:e2111477118. doi: 10.1073/pnas.2111477118. Available from: https://www.pnas.org/doi/10.1073/pnas.2111477118

30. Wagenhäuser I, Knies K, Hofmann D, Rauschenberger V, Eisenmann M, Reusch J, Gabel A, Flemming S, Andres O, Petri N, Topp MS, Papsdorf M, McDonogh M, Verma-Führing R, Scherzad A, Zeller D, Böhm H, Gesierich A, Seitz AK, Kiderlen M, Gawlik M, Taurines R, Wurmb T, Ernestus RI, Forster J, Weismann D, Weißbrich B, Dölken L, Liese J, Kaderali L, Kurzai O, Vogel U, and Krone M. Virus variant–specific clinical performance of SARS coronavirus two rapid antigen tests in point-of-care use, from November 2020 to January 2022. Clinical Microbiology and Infection 2023 Feb; 29:225–32. doi: 10.1016/j.cmi.2022.08.006. Available from: https://www.sciencedirect.com/science/article/pii/S1198743X22004220

31. Marc A, Kerioui M, Blanquart F, Bertrand J, Mitjà O, Corbacho-Monné M, Marks M, and Guedj J. Quantifying the relationship between SARS-CoV-2 viral load and infectiousness. eLife 2021 Sep; 10. Ed. by Cobey SE and Van der Meer JW. Publisher: eLife Sciences Publications, Ltd:e69302. doi: 10.7554/eLife.69302. Available from: 10.7554/eLife.69302

32. Brauner JM, Mindermann S, Sharma M, Johnston D, Salvatier J, Gavenčiak T, Stephenson AB, Leech G, Altman G, Mikulik V, Norman AJ, Monrad JT, Besiroglu T, Ge H, Hartwick MA, Teh YW, Chindelevitch L, Gal Y, and Kulveit J. Inferring the effectiveness of government interventions against COVID-19. Science 2021 Feb; 371. Publisher: American Association for the Advancement of Science:eabd9338. doi: 10.1126/science.abd9338. Available from: https://www.science.org/doi/full/10.1126/science.abd9338

33. Karimzadeh S, Bhopal R, and Nguyen Tien H. Review of infective dose, routes of transmission and outcome of COVID-19 caused by the SARS-COV-2: comparison with other respiratory viruses. Epidemiology and Infection 2021 Apr; 149:e96. doi: 10.1017/S0950268821000790. Available from: https://www.ncbi.nlm.nih.gov/pmc/articles/PMC8082124/

34. Aynaud MM, Hernandez JJ, Barutcu S, Braunschweig U, Chan K, Pearson JD, Trcka D, Prosser SL, Kim J, Barrios-Rodiles M, Jen M, Song S, Shen J, Bruce C, Hazlett B, Poutanen S, Attisano L, Bremner R, Blencowe BJ, Mazzulli T, Han H, Pelletier L, and Wrana JL. A multiplexed, next generation sequencing platform for high-throughput detection of SARS-CoV-2. Nature Communications 2021 Mar; 12. Number: 1 Publisher: Nature Publishing Group:1405. doi: 10.1038/s41467-021-21653-y. Available from: https://www.nature.com/articles/s41467-021-21653-y

35. Schreiber M. ‘We are not testing enough’: new US bird flu cases stoke fears over poor response. The Guardian 2024 Sep. Available from: https://www.theguardian.com/us-news/2024/sep/23/bird-flu-spreading-testing

36. Srimokla O and Petrie J. Effective contact tracing can halve mass testing frequency needed to control respiratory virus pandemics. 2023 Oct. doi: 10.1101/2023.10.12.23296969. Available from: https://www.medrxiv.org/content/10.1101/2023.10.12.23296969v1 [Accessed on: 2023 Nov 16]

37. Montazeri NX and Sandbrink JB. Innovate and Stockpile: Respiratory Protection for Essential Workers in a Catastrophic Pandemic. Health Security 2023 Aug; 21. Publisher: Mary Ann Liebert, Inc., publishers:266–71. doi: 10.1089/hs.2022.0126. Available from: https://www.liebertpub.com/doi/full/10.1089/hs.2022.0126

38. Gottberg C von, Krumm S, Porzsolt F, and Kilian R. The analysis of factors affecting municipal employees’ willingness to report to work during an influenza pandemic by means of the extended parallel process model (EPPM). BMC Public Health 2016 Jan; 16:26. doi: 10.1186/s12889-015-2663-8. Available from: 10.1186/s12889-015-2663-8

39. Hay JA, Hellewell J, and Qiu X. When intuition falters: repeated testing accuracy during an epidemic. European Journal of Epidemiology 2021 Jul; 36:749–52. doi: 10.1007/s10654-021-00786-w. Available from: 10.1007/s10654-021-00786-w

40. Han AX, Girdwood SJ, Khan S, Sacks JA, Toporowski A, Huq N, Hannay E, Russell CA, and Nichols BE. Strategies for Using Antigen Rapid Diagnostic Tests to Reduce Transmission of Severe Acute Respiratory Syndrome Coronavirus 2 in Low- and Middle-Income Countries: A Mathematical Modelling Study Applied to Zambia. Clinical Infectious Diseases 2023 Feb; 76:620–30. doi: 10.1093/cid/ciac814. Available from: 10.1093/cid/ciac814

41. Cutler DM and Summers LH. The COVID-19 Pandemic and the $16 Trillion Virus. JAMA 2020 Oct; 324:1495–6. doi: 10.1001/jama.2020.19759. Available from: 10.1001/jama.2020.19759

42. Psacharopoulos G, Patrinos H, Collis V, and Vegas E. The COVID-19 cost of school closures. 2020 Apr. Available from: https://www.brookings.edu/articles/the-covid-19-cost-of-school-closures/ [Accessed on: 2023 Aug 15]

43. Guglielmi G. Rapid coronavirus tests: a guide for the perplexed. Nature 2021 Feb; 590. Bandiera abtest: a Cg type: News Feature Number: 7845 Publisher: Nature Publishing Group Subject term: SARS-CoV-2, Virology, Immunology:202–5. doi: 10.1038/d41586-021-00332-4. Available from: https://www.nature.com/articles/d41586-021-00332-4

44. Gu W, Deng X, Lee M, Sucu YD, Arevalo S, Stryke D, Federman S, Gopez A, Reyes K, Zorn K, Sample H, Yu G, Ishpuniani G, Briggs B, Chow ED, Berger A, Wilson MR, Wang C, Hsu E, Miller S, DeRisi JL, and Chiu CY. Rapid pathogen detection by metagenomic next-generation sequencing of infected body fluids. Nature Medicine 2021 Jan; 27. Number: 1 Publisher: Nature Publishing Group:115–24. doi: 10.1038/s41591-020-1105-z. Available from: https://www.nature.com/articles/s41591-020-1105-z

45. Lu R, Wu X, Wan Z, Li Y, Jin X, and Zhang C. A Novel Reverse Transcription Loop-Mediated Isothermal Amplification Method for Rapid Detection of SARS-CoV-2. International Journal of Molecular Sciences 2020 Apr; 21:2826. doi: 10.3390/ijms21082826

46. Broughton JP, Deng X, Yu G, Fasching CL, Servellita V, Singh J, Miao X, Streithorst JA, Granados A, Sotomayor-Gonzalez A, Zorn K, Gopez A, Hsu E, Gu W, Miller S, Pan CY, Guevara H, Wadford DA, Chen JS, and Chiu CY. CRISPR–Cas12-based detection of SARS-CoV-2. Nature Biotechnology 2020 Jul; 38. Number: 7 Publisher: Nature Publishing Group:870–4. doi: 10.1038/s41587-020-0513-4. Available from: https://www.nature.com/articles/s41587-020-0513-4

47. Shiaelis N, Tometzki A, Peto L, McMahon A, Hepp C, Bickerton E, Favard C, Muriaux D, Andersson M, Oakley S, Vaughan A, Matthews PC, Stoesser N, Crook DW, Kapanidis AN, and Robb NC. Virus Detection and Identification in Minutes Using Single-Particle Imaging and Deep Learning. ACS Nano 2023 Jan; 17. Publisher: American Chemical Society:697–710. doi: 10.1021/acsnano.2c10159. Available from: 10.1021/acsnano.2c10159

48. Corman VM, Landt O, Kaiser M, Molenkamp R, Meijer A, Chu DK, Bleicker T, Brünink S, Schneider J, Schmidt ML, Mulders DG, Haagmans BL, Veer B van der, Brink S van den, Wijsman L, Goderski G, Romette JL, Ellis J, Zambon M, Peiris M, Goossens H, Reusken C, Koopmans MP, and Drosten C. Detection of 2019 novel coronavirus (2019-nCoV) by real-time RT-PCR. Eurosurveillance 2020 Jan; 25:2000045. doi: 10.2807/1560-7917.ES.2020.25.3.2000045. Available from: https://www.ncbi.nlm.nih.gov/pmc/articles/PMC6988269/

49. Li D, Zhang J, and Li J. Primer design for quantitative real-time PCR for the emerging Coronavirus SARS-CoV-2. Theranostics 2020 Jun; 10:7150–62. doi: 10.7150/thno.47649. Available from: https://www.ncbi.nlm.nih.gov/pmc/articles/PMC7330846/

50. Jarvis B, Pavone T, and Cedillo I. Designing Commercial-Scale Oligonucleotide Synthesis. Pharmaceutical Technology. Pharmaceutical Technology-02-02-2020 2020 Feb; 44. Publisher: MJH Life Sciences:30–4. Available from: https://www.pharmtech.com/view/designing-commercial-scale-oligonucleotide-synthesis

51. Why the CDC botched its coronavirus testing. Available from: https://www.technologyreview.com/2020/03/05/905484/why-the-cdc-botched-its-coronavirus-testing/ [Accessed on: 2023 Aug 10]

52. Tobik ER, Kitfield-Vernon LB, Thomas RJ, Steel SA, Tan SH, Allicock OM, Choate BL, Akbarzada S, and Wyllie AL. Saliva as a sample type for SARS-CoV-2 detection: implementation successes and opportunities around the globe. Expert Review of Molecular Diagnostics 2022 May; 22. Publisher: Taylor & Francis eprint: 10.1080/14737159.2022.2094250:519-35. doi: 10.1080/14737159.2022.2094250. Available from: 10.1080/14737159.2022.2094250

53. Bastos ML, Perlman-Arrow S, Menzies D, and Campbell JR. The Sensitivity and Costs of Testing for SARS-CoV-2 Infection With Saliva Versus Nasopharyngeal Swabs. Annals of Internal Medicine 2021 Apr; 174. Publisher: American College of Physicians:501–10. doi: 10.7326/M20-6569. Available from: https://www.acpjournals.org/doi/full/10.7326/M20-6569

54. Vogels CBF, Watkins AE, Harden CA, Brackney DE, Shafer J, Wang J, Caraballo C, Kalinich CC, Ott IM, Fauver JR, Kudo E, Lu P, Venkataraman A, Tokuyama M, Moore AJ, Muenker MC, Casanovas-Massana A, Fournier J, Bermejo S, Campbell M, Datta R, Nelson A, Anastasio K, Askenase MH, Batsu M, Bickerton S, Brower K, Bucklin ML, Cahill S, Cao Y, Courchaine E, DeIuliis G, Earnest R, Geng B, Goldman-Israelow B, Handoko R, Khoury-Hanold W, Kim D, Knaggs L, Kuang M, Lapidus S, Lim J, Linehan M, Lu-Culligan A, Martin A, Matos I, McDonald D, Minasyan M, Nakahata M, Naushad N, Nouws J, Obaid A, Odio C, Oh JE, Omer S, Park A, Park HJ, Peng X, Petrone M, Prophet S, Rice T, Rose KA, Sewanan L, Sharma L, Shaw AC, Shepard D, Smolgovsky M, Sonnert N, Strong Y, Todeasa C, Valdez J, Velazquez S, Vijayakumar P, White EB, Yang Y, Dela Cruz CS, Ko AI, Iwasaki A, Krumholz HM, Matheus JD, Hui P, Liu C, Farhadian SF, Sikka R, Wyllie AL, and Grubaugh ND. SalivaDirect: A simplified and flexible platform to enhance SARS-CoV-2 testing capacity. Med 2021 Mar; 2:263–280.e6. doi: 10.1016/j.medj.2020.12.010. Available from: https://www.sciencedirect.com/science/article/pii/S2666634020300763

55. Cardwell K, O’Neill SM, Tyner B, Broderick N, O’Brien K, Smith SM, Harrington P, Ryan M, and O’Neill M. A rapid review of measures to support people in isolation or quarantine during the Covid-19 pandemic and the effectiveness of such measures. Reviews in Medical Virology 2022; 32. eprint: https://onlinelibrary.wiley.com/doi/pdf/10.1002/rmv.2244:e2244. doi: 10.1002/rmv.2244. Available from: https://onlinelibrary.wiley.com/doi/abs/10.1002/rmv.2244

56. Grout L, Katar A, Ait Ouakrim D, Summers JA, Kvalsvig A, Baker MG, Blakely T, and Wilson N. Failures of quarantine systems for preventing COVID-19 outbreaks in Australia and New Zealand. Medical Journal of Australia 2021; 215:320–4. doi: 10.5694/mja2.51240. Available from: https://onlinelibrary.wiley.com/doi/abs/10.5694/mja2.51240

57. DiGiovanni C, Conley J, Chiu D, and Zaborski J. Factors Influencing Compliance with Quarantine in Toronto During the 2003 SARS Outbreak. Biosecurity and Bioterrorism: Biodefense Strategy, Practice, and Science 2004 Dec; 2. Publisher: Mary Ann Liebert, Inc., publishers:265–72. doi: 10.1089/bsp.2004.2.265. Available from: https://www.liebertpub.com/doi/10.1089/bsp.2004.2.265

58. Li Z, Liu F, Cui J, Peng Z, Chang Z, Lai S, Chen Q, Wang L, Gao GF, and Feng Z. Comprehensive large-scale nucleic acid–testing strategies support China’s sustained containment of COVID-19. Nature Medicine 2021 May; 27. Number: 5 Publisher: Nature Publishing Group:740–2. doi: 10.1038/s41591-021-01308-7. Available from: https://www.nature.com/articles/s41591-021-01308-7

59. Minhas N, Gurav YK, Sambhare S, Potdar V, Choudhary ML, Bhardwaj SD, and Abraham P. Cost-analysis of real time RT-PCR test performed for COVID-19 diagnosis at India’s national reference laboratory during the early stages of pandemic mitigation. PLOS ONE 2023 Jan; 18. Publisher: Public Library of Science:e0277867. doi: 10.1371/journal.pone.0277867. Available from: https://journals.plos.org/plosone/article?id=10.1371/journal.pone.0277867

60. Home Office. Fire and rescue workforce and pensions statistics: England, April 2021 to March 2022. Available from: https://www.gov.uk/government/statistics/fire-and-rescue-workforce-and-pensions-statistics-england-april-2021-to-march-2022/fire-and-rescue-workforce-and-pensions-statistics-england-april-2021-to-march-2022 [Accessed on: 2023 Aug 12]

61. Diagnostic & Medical Laboratories in the US - Market Size, Industry Analysis, Trends and Forecasts (2023-2028)— IBISWorld. Available from: https://www.ibisworld.com/default.aspx [Accessed on: 2023 Nov 2]

62. Landaverde L, McIntyre D, Robson J, Fu D, Ortiz L, Chen R, Oliveira SMD, Fan A, Barrett A, Burgay SP, Choate S, Corbett D, Doucette-Stamm L, Gonzales K, Hamer DH, Huang L, Huval S, Knight C, Landa C, Lindquist D, Lockard K, Macdowell TL, Mauro E, McGinty C, Miller C, Monahan M, Moore R, Platt J, Rolles L, Roy J, Schroeder T, Tolan DR, Zaia A, Brown RA, Waters G, Densmore D, and Klapperich CM. Buildout and integration of an automated high-throughput CLIA laboratory for SARS-CoV-2 testing on a large urban campus. SLAS Technology 2022 Oct; 27. Publisher: Elsevier:302–11. doi: 10.1016/j.slast.2022.06.003. Available from: https://slas-technology.org/article/S2472-6303(22)05163-9/fulltext

63. Douthwaite JA, Brown CA, Ferdinand JR, Sharma R, Elliott J, Taylor MA, Malintan NT, Duvoisin H, Hill T, Delpuech O, Orton AL, Pitt H, Kuenzi F, Fish S, Nicholls DJ, Cuthbert A, Richards I, Ratcliffe G, Upadhyay A, Marklew A, Hewitt C, Ross-Thriepland D, Brankin C, Chodorge M, Browne G, Mander PK, DeWildt RM, Weaver S, Smee PA, Kempen J van, Bartlett JG, Allen PM, Koppe EL, Ashby CA, Phipps JD, Mehta N, Brierley DJ, Tew DG, Leveridge MV, Baddeley SM, Goodfellow IG, Green C, Abell C, Neely A, Waddell I, Rees S, Maxwell PH, Pangalos MN, Howes R, and Clark R. Improving the efficiency and effectiveness of an industrial SARS-CoV-2 diagnostic facility. Scientific Reports 2022 Feb; 12. Number: 1 Publisher: Nature Publishing Group:3114. doi: 10.1038/s41598-022-06873-6. Available from: https://www.nature.com/articles/s41598-022-06873-6

64. Krijger PHL, Hoek TA, Boersma S, Donders LIPM, Broeders MMC, Pieterse M, Toonen PW, Logister I, Verhagen BMP, Verstegen MJAM, Ravesteyn TW van, Roymans RJTM, Mattiroli F, Vandesompele J, Nijhuis M, Meijer S, Weert A van, Dekker E, Dom FJ, Ruijtenbeek R, Velden LBJ van der, Bovenkamp JHB van de, Bosch M, Laat W de, and Tanenbaum ME. A public–private partnership model for COVID-19 diagnostics. Nature Biotechnology 2021 Oct; 39. Number: 10 Publisher: Nature Publishing Group:1182–4. doi: 10.1038/s41587-021-01080-6. Available from: https://www.nature.com/articles/s41587-021-01080-6

65. Moses S, Warren C, Robinson P, Curtis J, Asquith S, Holme J, Jain N, Brookes KJ, and Hanley QS. Endpoint PCR Detection of Sars-CoV-2 RNA. 2020 Jul. doi: 10.1101/2020.07.21.20158337. Available from: https://www.medrxiv.org/content/10.1101/2020.07.21.20158337v1 [Accessed on: 2023 Aug 12]

66. Ultra-high-throughput PCR testing system for SARS-CoV-2 detection. Available from: https://www.biosearchtech.com/ultra-high-throughput-pcr-testing-system-for-sars-cov-2-detection [Accessed on: 2023 Aug 12]

67. SUMMARY NOTE: VISIT TO THE ROSALIND FRANKLIN LABORATORY, LEAMINGTON SPA (DECEMBER 2021). Available from: https://committees.parliament.uk/writtenevidence/107131/html/ [Accessed on: 2023 Aug 12]

68. Lovett S. Inside Britain’s flagship Covid lab that no one knows what to do with. 2022 Apr. Available from: https://www.independent.co.uk/news/health/covid-test-rosalind-franklin-laboratory-b2067802.html [Accessed on: 2023 Aug 12]

69. Cost of processing a PCR test for Covid-19: FOI release. Available from: http://www.gov.scot/publications/foi-202100215784/ [Accessed on: 2023 Aug 12]

70. Mendoza RP, Bi C, Cheng HT, Gabutan E, Pagaspas GJ, Khan N, Hoxie H, Hanna S, Holmes K, Gao N, Lewis R, Wang H, Neumann D, Chan A, Takizawa M, Lowe J, Chen X, Kelly B, Asif A, Barnes K, Khan N, May B, Chowdhury T, Pollonini G, Gouda N, Guy C, Gordon C, Ayoluwa N, Colon E, Miller-Medzon N, Jones S, Hossain R, Dodson A, Weng M, McGaskey M, Vasileva A, Lincoln AE, Sikka R, Wyllie AL, Berke EM, Libien J, Pincus M, and Premsrirut PK. Implementation of a pooled surveillance testing program for asymptomatic SARS-CoV-2 infections in K-12 schools and universities. eClinicalMedicine 2021 Aug; 38. Publisher: Elsevier. doi: 10.1016/j.eclinm.2021.101028. Available from: https://www.thelancet.com/journals/eclinm/article/PIIS2589-5370(21)00308-4/fulltext#%20

71. Taylor A, Fram A, Kellman L, and Superville D. Trump signs $2.2T stimulus after swift congressional votes. 2020 Mar. Available from: https://apnews.com/article/donald-trump-financial-markets-ap-top-news-bills-virus-outbreak-2099a53bb8adf2def7ee7329ea322f9d [Accessed on: 2023 Aug 12]

